# Equality in Hearing Aid Access: A Systematic Review and Meta-analysis

**DOI:** 10.64898/2025.12.03.25341553

**Authors:** Esther K. Hui, Naaheed Mukadam, Zuyu Wang, Emanuelle Rossetti, Louise Marston, Gill Livingston

**Affiliations:** Department of Mental Health of Older People, Division of Psychiatry, University College London; North London NHS Foundation Trust, London; Department of Primary Care and Population Health, University College London

## Abstract

**Importance:** Most people with hearing impairment do not acquire hearing aids, but it is unknown what sociodemographic factors influence access.

**Objective:** To systematically review, synthesize and meta-analyze sociodemographic characteristics of people who do and do not acquire hearing aids after hearing loss diagnosis.

**Data sources:** We pre-registered the study (PROSPERO: CRD42023428580), and searched MEDLINE, EMBASE, and Web of Science from inception to 27 October 2025. Search terms included hearing aids and cohort study terms.

**Study selection:** We included observational studies with participants aged ≥18 years who had hearing loss confirmed through pure-tone audiometry, with any sociodemographic characteristics of those who do and do not access hearing aids.

**Data extraction and synthesis:** Using the Meta-analysis of Observational Studies in Epidemiology (MOOSE) guidelines, three authors independently extracted data, assessing study quality using the Newcastle-Ottawa Scale. We calculated ratios of people with hearing loss from each sociodemographic characteristic who acquired hearing aids. We used random effects meta-analyses.

**Main outcomes and measures:** Hearing aid acquisition.

**Results:** 36 studies including 300,946 people met criteria. Men were more likely to acquire hearing aids than women (n, 287,964; RR, 1.09[1.02-1.17]; I^2^, 98.7%), White people more than other ethnic/racial groups (n, 274,860; RR, 1.26[1.07-1.55]; I^2^, 99.9%), >12 years vs. ≤12 years of education (n, 5,970; RR, 1.17[1.04, 1.32]; I^2^, 79.4%), and pension recipients more than non-recipients (n, 1,678; RR, 1.50 [1.08,2.09], I^2^, 87.4%). There were no differences in hearing aid acquisition by very low household income (≥US$45,000/year vs. <US$45,000/year; RR, 1.06[0.87-1.30]; I^2^, 0%), employment status (currently employed vs. not employed (RR, 0.58[0.34-1.00]; I^2^, 84.2%), relationship status (currently married/partnered vs. not; RR, 0.96[0.87-1.16]; I^2^, 70.0%), living alone vs. with others (RR, 0.82[0.57-1.18]; I^2^, 97.7%) or rural vs. urban areas (RR, 1.20[0.86-1.68]; I^2^, 94.4%).

**Conclusion and Relevance:** Underserved groups (women, minorities, those with less education and not receiving pensions) are less likely to get hearing aids, even after hearing testing.

Meta-analyses had high heterogeneity, so findings are not generalizable. Some underserved people can access hearing aids, and it is important to further investigate the barriers and enablers. No studies controlled for hearing severity, so findings are limited by confounding by indication.

**Key Points:** *Question:* What groups of people with diagnosed hearing loss acquire hearing aids? Finding: Among 300,946 individuals with hearing loss (36 studies), men, those with >12 years of education, pension recipients, and White individuals were more likely to acquire hearing aids. We found no differences based on household income, employment status, relationship status, rural vs. urban areas, and living arrangement.

*Meaning:* After accessing hearing services, hearing aid acquisition is related to sex, ethnicity or race, education, and pension status, but no other sociodemographic indicators. Further research is needed about enablers and barriers to obtaining hearing aids for women and ethnic minorities.

## Introduction

The World Health Organization (WHO) estimates that more than 1.5 billion people, or 20% of the global population, currently have hearing loss.^1^ One in four people globally are projected to have hearing impairment by 2050.^2^ Untreated or undiagnosed hearing loss not only has negative health consequences but also a significant impact on social, occupational, cognitive and emotional well-being.^3,4^ Sixty-eight percent of people with hearing loss felt isolated at work because of their impairment.^5^ Having hearing loss doubles the risk of developing depression, ^5^ and is associated with the risk of dementia.^4,6^ However, using hearing aids can mitigate some of the negative impacts. For example, a meta-analysis of 31 studies of people with hearing loss reported hearing restorative devices are linked to a 19% decrease in long-term cognitive decline and a 3% improvement in cognitive test scores.^7^ Similarly, a study of 17,000 participants from 28 countries found hearing aid use was associated with lower rates of depression.^8^

Despite the benefits of hearing aids and variation in ownership and usage among different populations, fewer than half have them, and only half use them most of the time.^9,10^ Hearing aid access may be hindered by factors,^11^ such as living in rural areas, low household income, stigma from wearing hearing aids, and having comorbidities, such as diabetes, hypertension, and a history of stroke. ^3,12^ People from lower- and middle-income countries, as well as from minority ethnic and underserved groups, are found to be less likely to access hearing aids.^3,11,13^ There are conflicting findings about age and sex. ^11,13,14^

With one narrative review finding they did not impact uptake,^14^ but a systematic review found males were more affected by the stigma of wearing hearing aids, and the degree of hearing loss influenced females to get hearing aids.^13^ This review did not explain to what degree age and sex impact hearing aid uptake, or meta-analysed sociodemographic factors. and ^11,13,14^

This review aims to systematically review, synthesize and meta-analyze the personal characteristics of people who acquire and do not acquire hearing aids—to our knowledge, for the first time. This is to help understand the factors that affect access to hearing aids, to inform future interventions and improve access. We hypothesize that underserved groups will access hearing aids less than those without these characteristics.

## Methods

We conducted this pre-registered study (PROSPERO: CRD42023428580) in accordance with the Meta-analysis of Observational Studies in Epidemiology (MOOSE) guidelines (eTable 1 Supplemental 1).

### Search strategies

We searched MEDLINE, EMBASE, and Web of Science, and used Covidence to organise the data.^15^ We restricted the search to human studies with no limits on language or date of publication. We used keywords and database-specific subject headings, developed in collaboration with a university librarian. GL and NM reviewed the search terms. We searched the title and abstract, using MeSH terms for each database covering “hearing aid” and “cohort studies.” EH conducted the initial search on 23 June 2024 and an updated search on 27 October 2025 (full search in eTable 2 Supplemental 1). We also searched the reference lists of previous systematic reviews and of included papers.

### Inclusion criteria

- Study design: observational studies.
- Participants: aged ≥18years who have hearing loss confirmed through pure-tone audiometry (PTA), as this is the gold standard clinical test for assessing hearing.
- Relevant exposure reported: at least one sociodemographic characteristic of people with hearing loss, i.e., sex, socioeconomic status, education, race/ethnicity, income.
- Comparisons: people without these characteristics.
- Outcomes: hearing aid (yes/no).

### Exclusion criteria

- People without hearing loss.

### Selection process and data extraction

ER, EH and ZW screened titles and abstracts and obtained the full-text articles. EH, ER and ZW independently assessed the full-text articles against the inclusion criteria. GL and NM resolved disagreements between the reviewers. We recorded the reasons for excluding studies. If the study had missing information, we contacted the authors for further details.

EH, ER, and ZW independently extracted relevant information from the full-text articles and discussed it among themselves when there were conflicting interpretations:

- Methodological characteristics: study design, country, number of participants, name of the trial.
- Participant characteristics reported: sex, age, education level (level and year of education), socioeconomic status, household income, race/ethnicity, relationship status, living alone or with others, urban or rural, insurance coverage, pension status, and current employment status.
- outcome: hearing aid access (yes/no).

For cohort and case-control studies, we only extracted cross-sectional data. We contacted authors of the included studies for any relevant data not reported in the paper.

### Quality assessment

Two reviewers, EH and ZW, independently appraised the included papers using the Newcastle Ottawa Criteria for cross-sectional studies.^16^ Some questions in the quality assessment were not applicable, such as “assessment of outcome” and “statistical test” items, as hearing aid access was self-reported and no statistical tests were applicable, so these questions were removed. For “ascertainment of exposure”, all included studies scored one because sociodemographic information was the exposure. For the “representativeness”, studies scored two if all participants were from a hearing clinic, and one if the cohort was focused on people with hearing loss. We gave a score of one to studies with 20% for “non-respondent”, and one for “comparability” if they reported demographic information categorising hearing severity by different groups. The possible total was five, with higher scores meaning better quality. EH and GL resolved discrepancies between the reviews and identified the reasons.

### Analysis

We described the findings narratively. We calculated the rate of a specific sociodemographic characteristic (e.g., sex) among individuals with hearing loss who acquired hearing aids compared to those with hearing loss who did not, to obtain a risk ratio (RR). Then, we pooled results from two or more studies which reported the same characteristics using the random effects model on R Studio (version 4.3.2). We evaluated the heterogeneity in meta-analysis using the I² statistic, which measures the proportion of variability attributed to differences in treatment effects beyond what would be expected by chance in a random-effects meta-analysis, with I² value between 75% and 100% indicating substantial heterogeneity.^17^

## Results

### Study selection

36/6,533 studies in the search fulfilled inclusion criteria. This includes five additional papers from previous systematic reviews, and eight from the updated search (see PRISMA diagram; Figure 1) (Table 1).^13^

**Figure 1.**
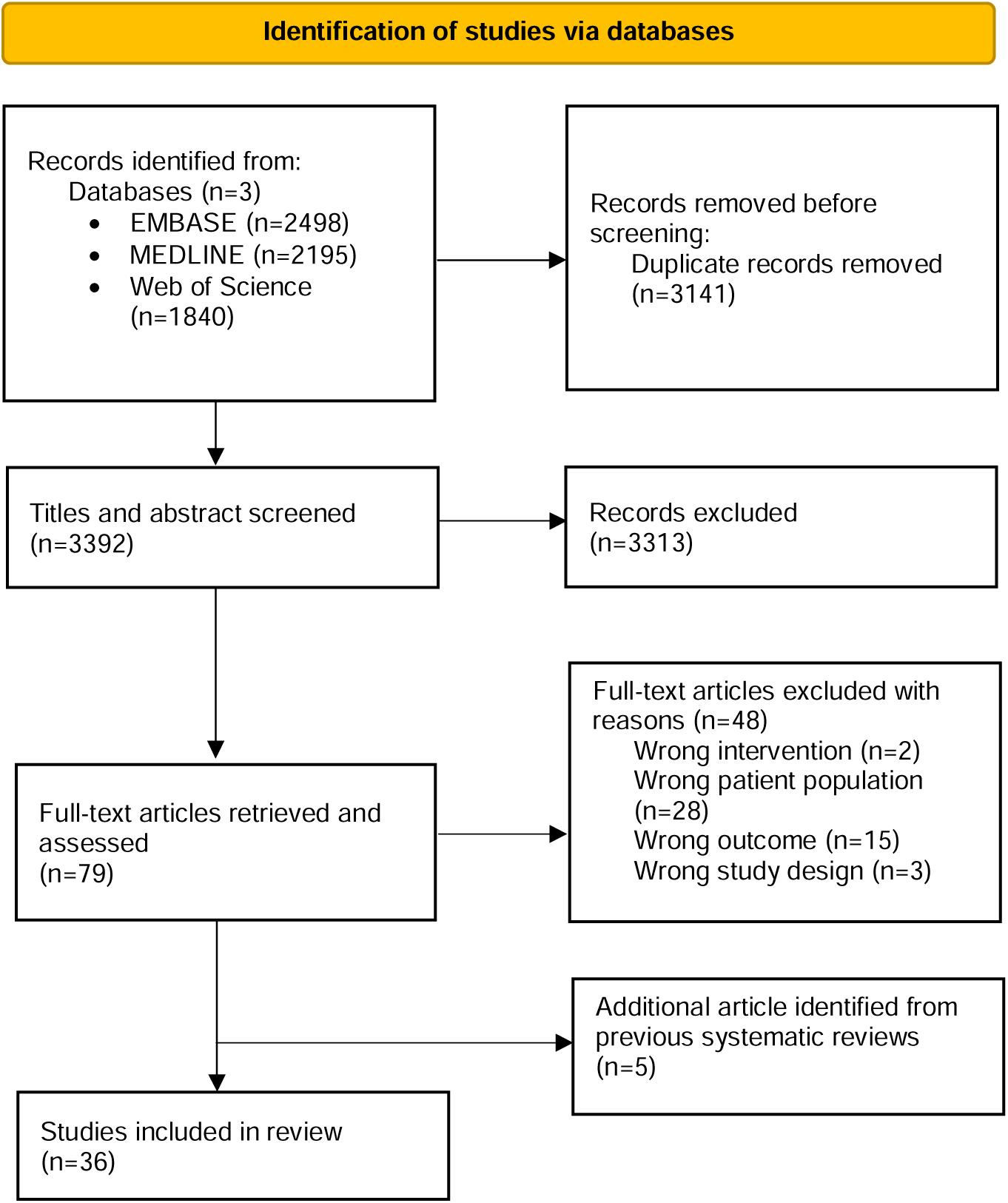
PRISMA diagram of study identification and selection.

**Table 1.**
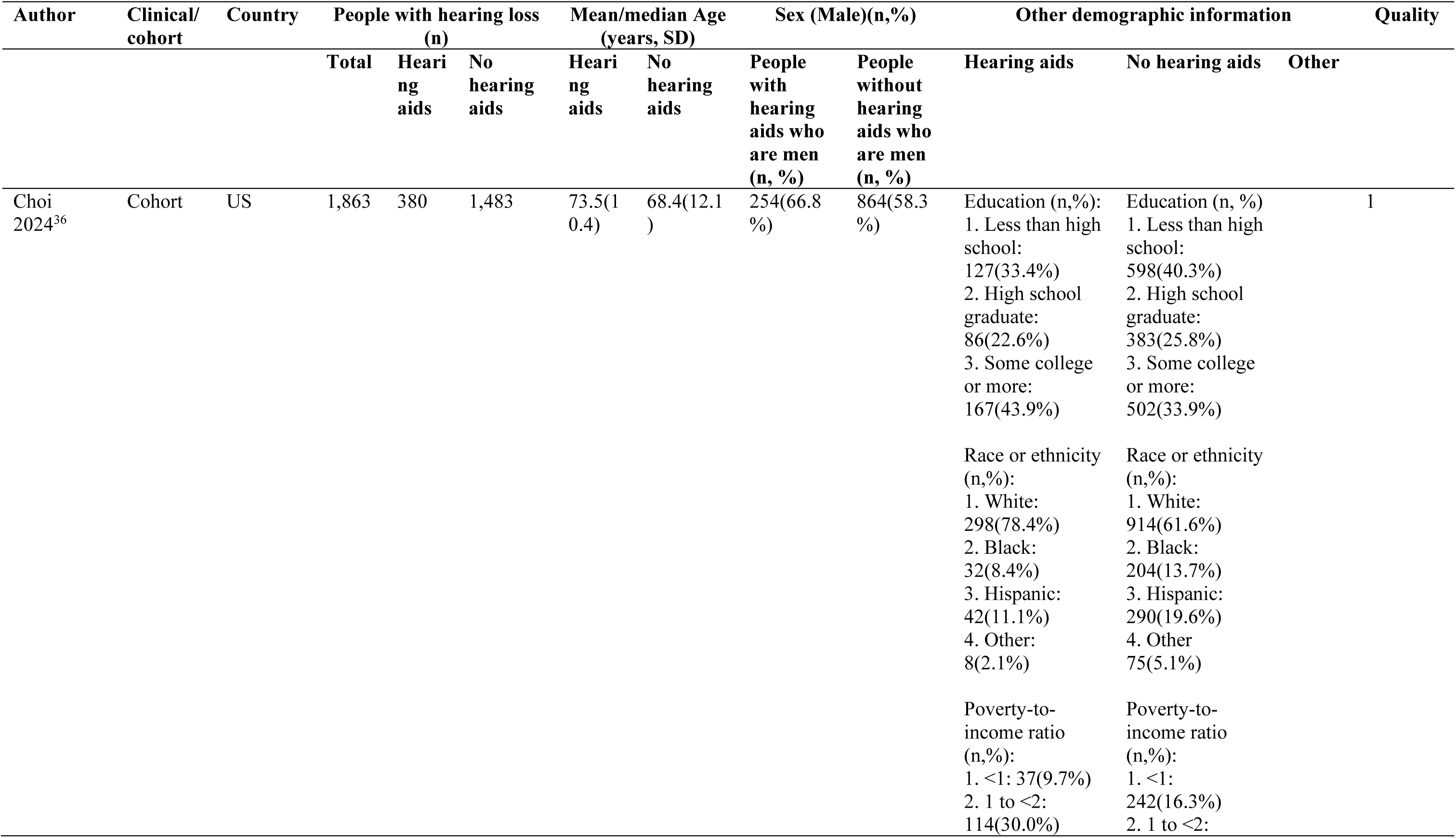
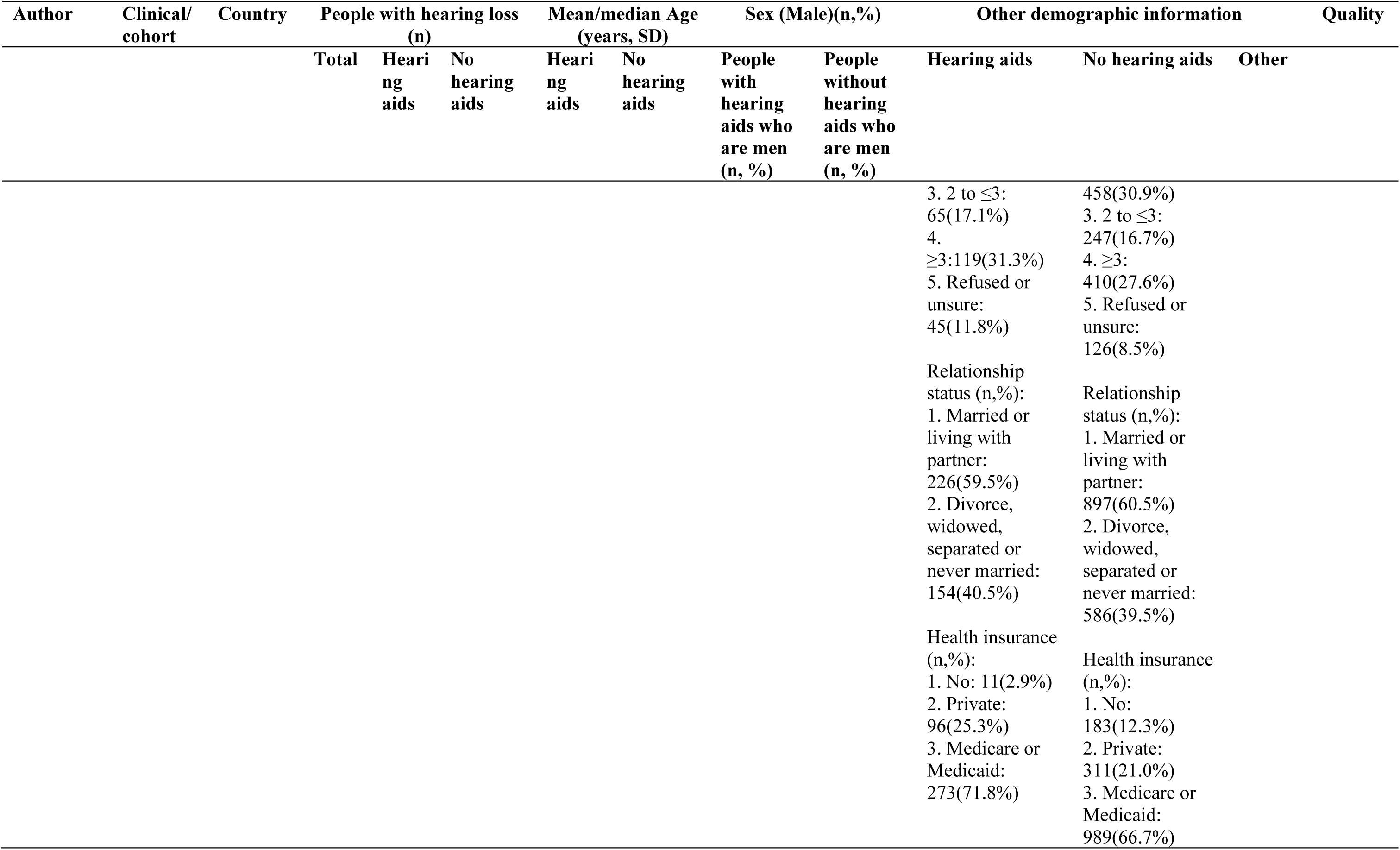
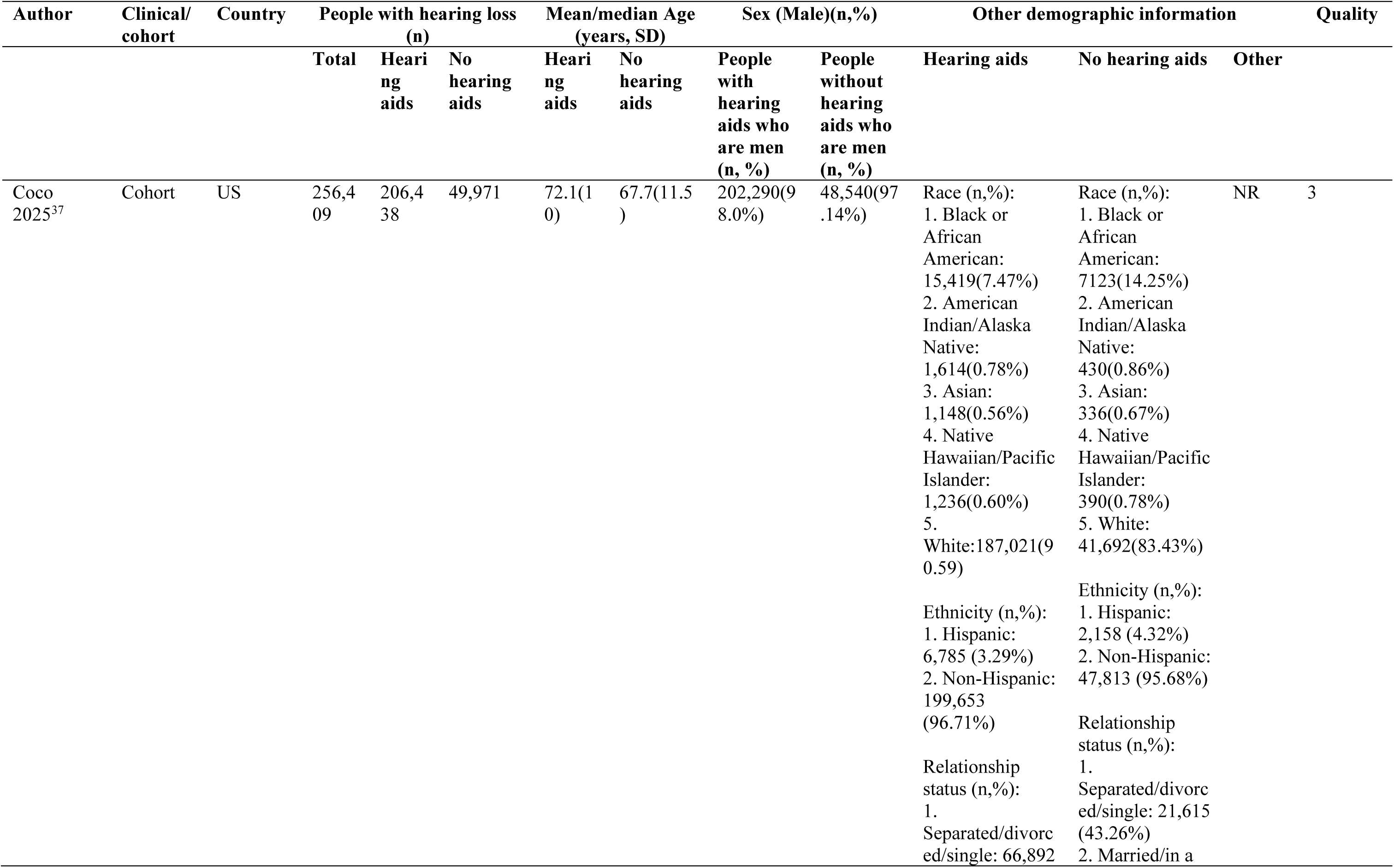
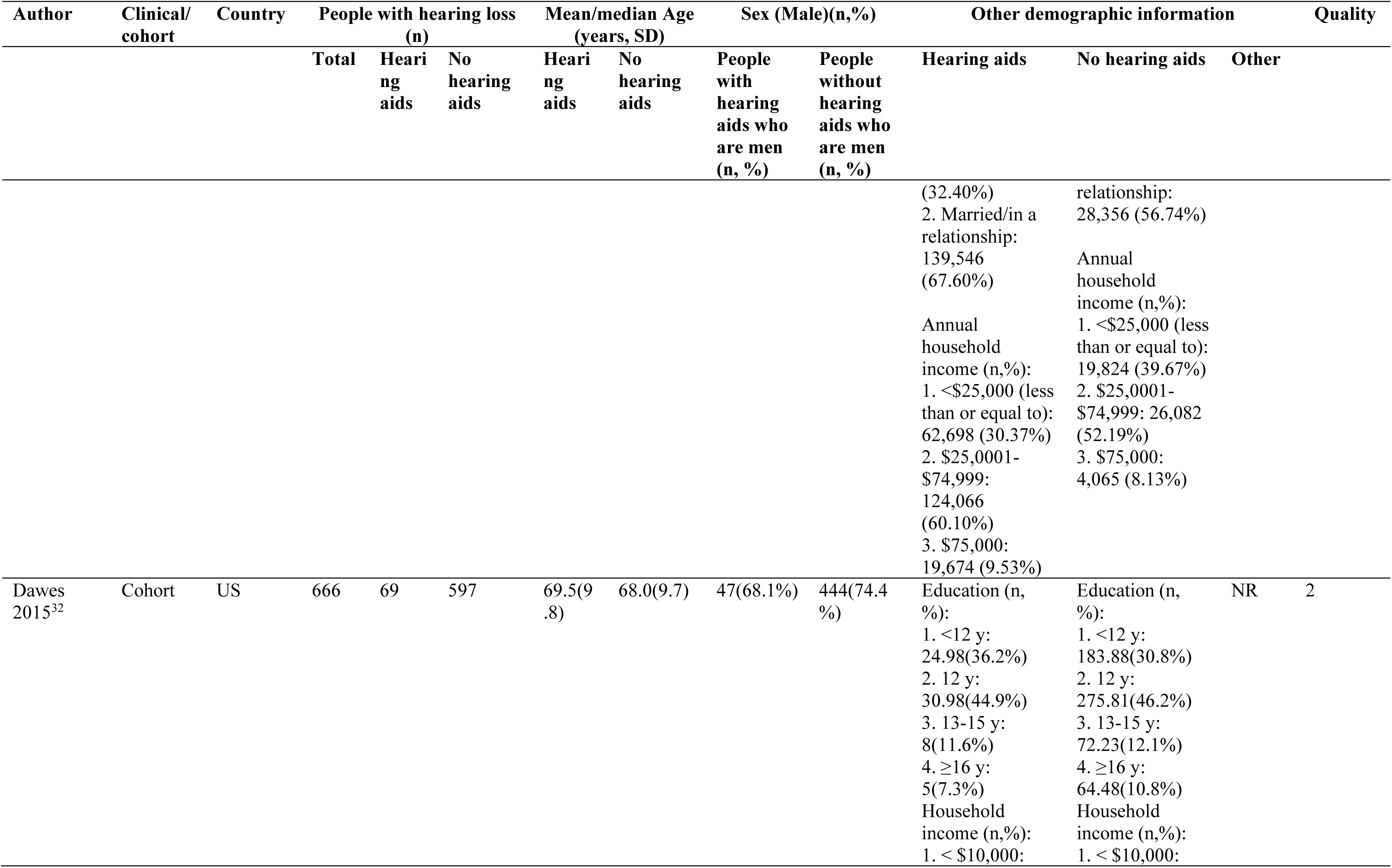
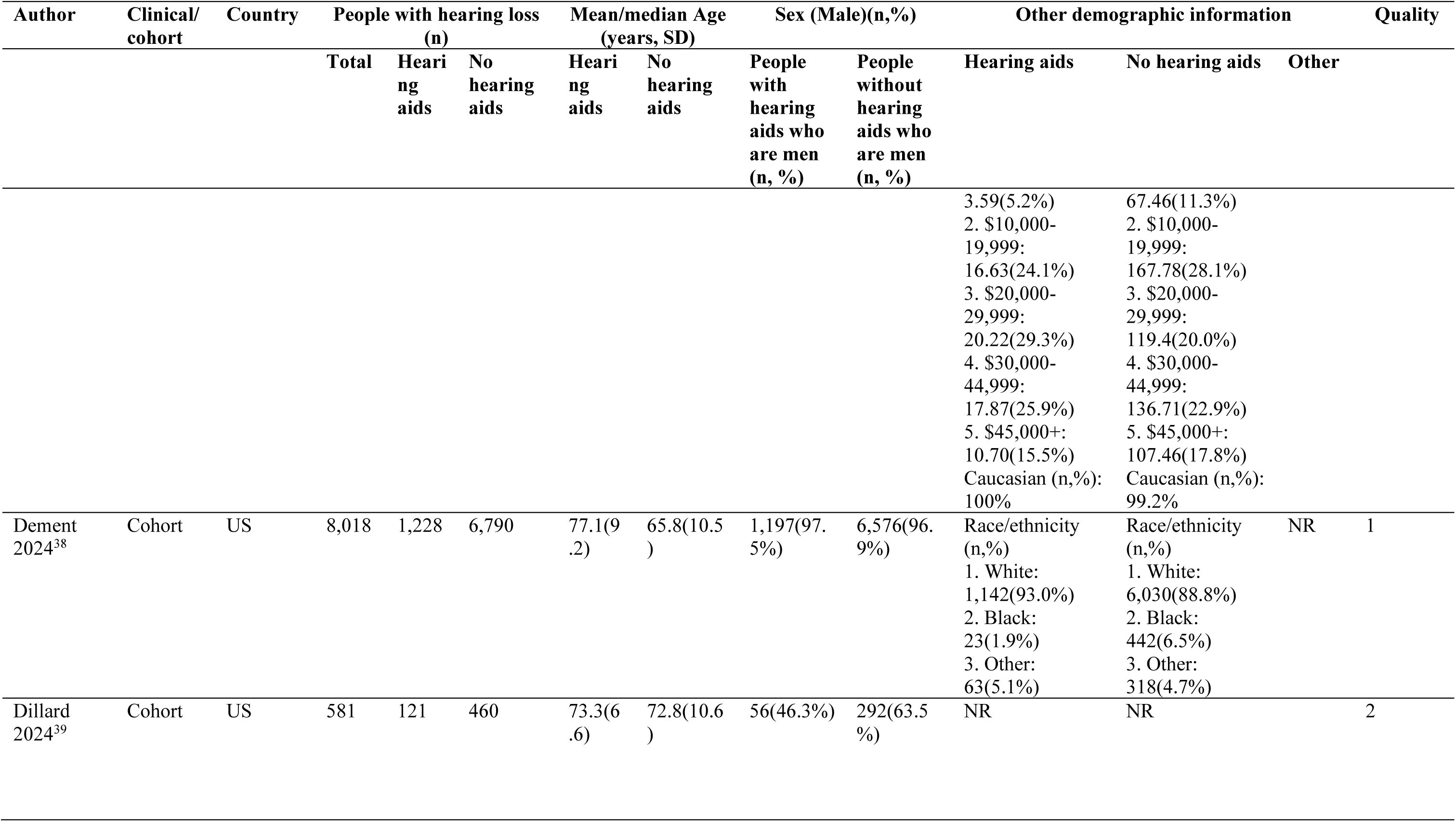
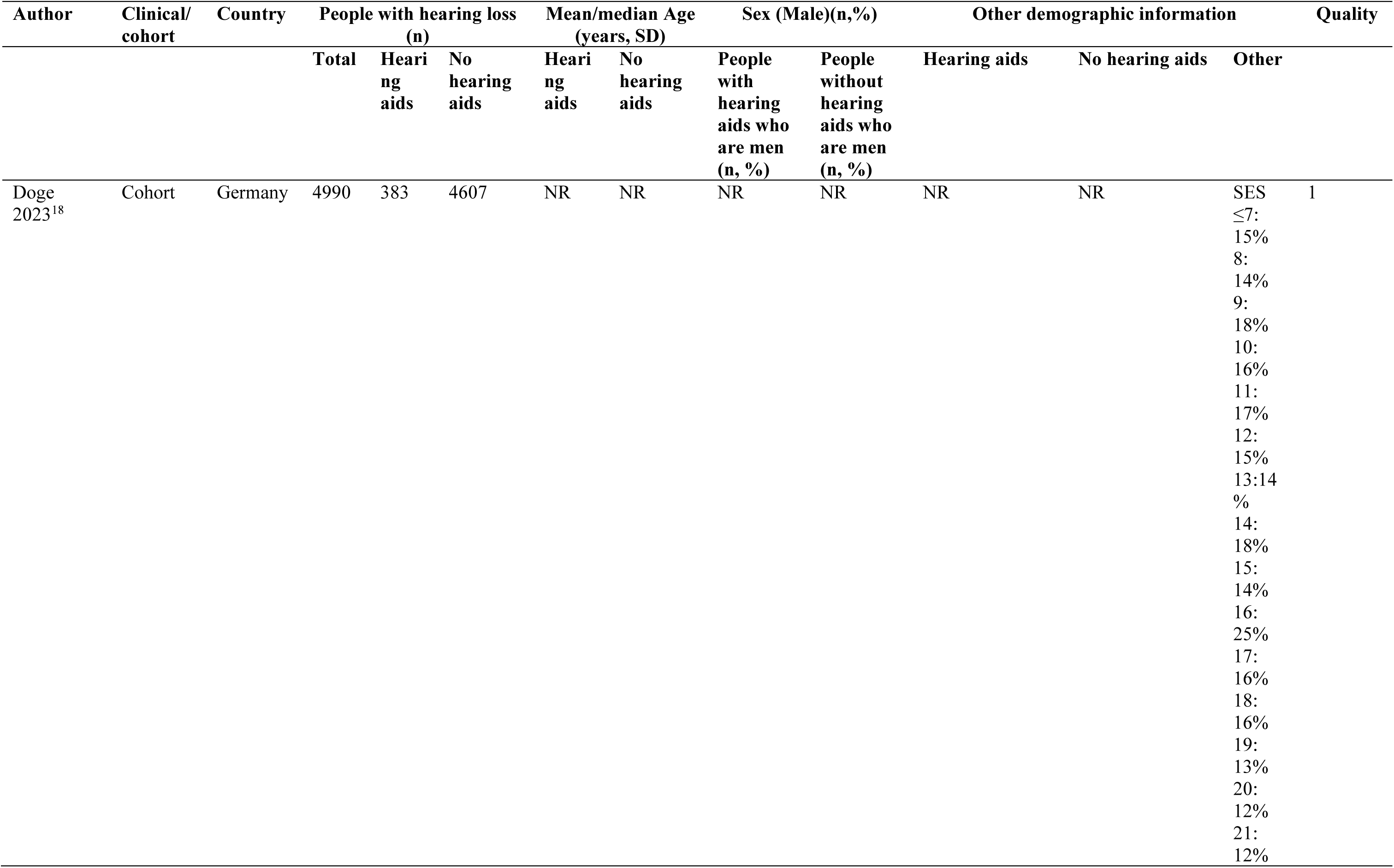
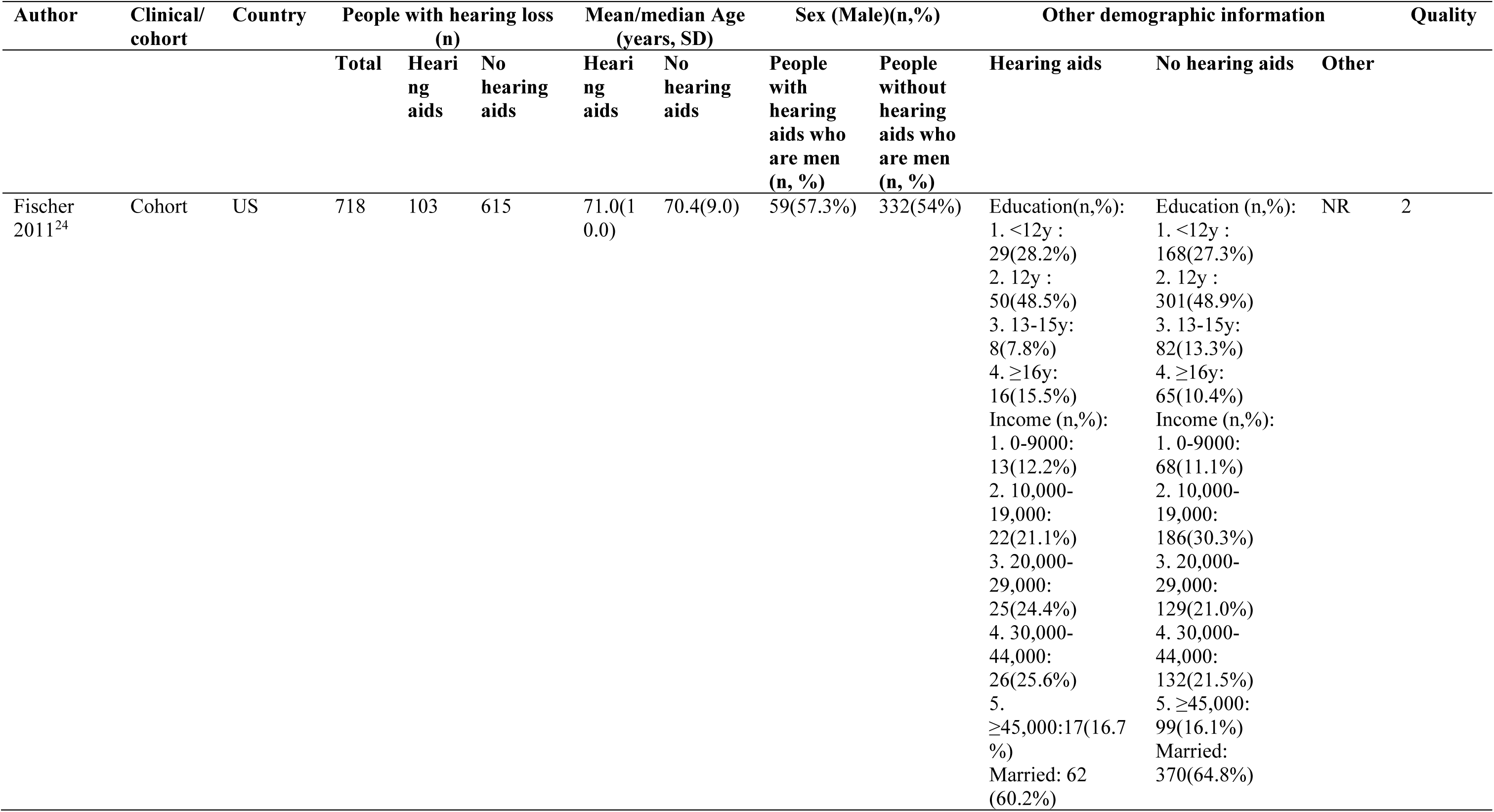
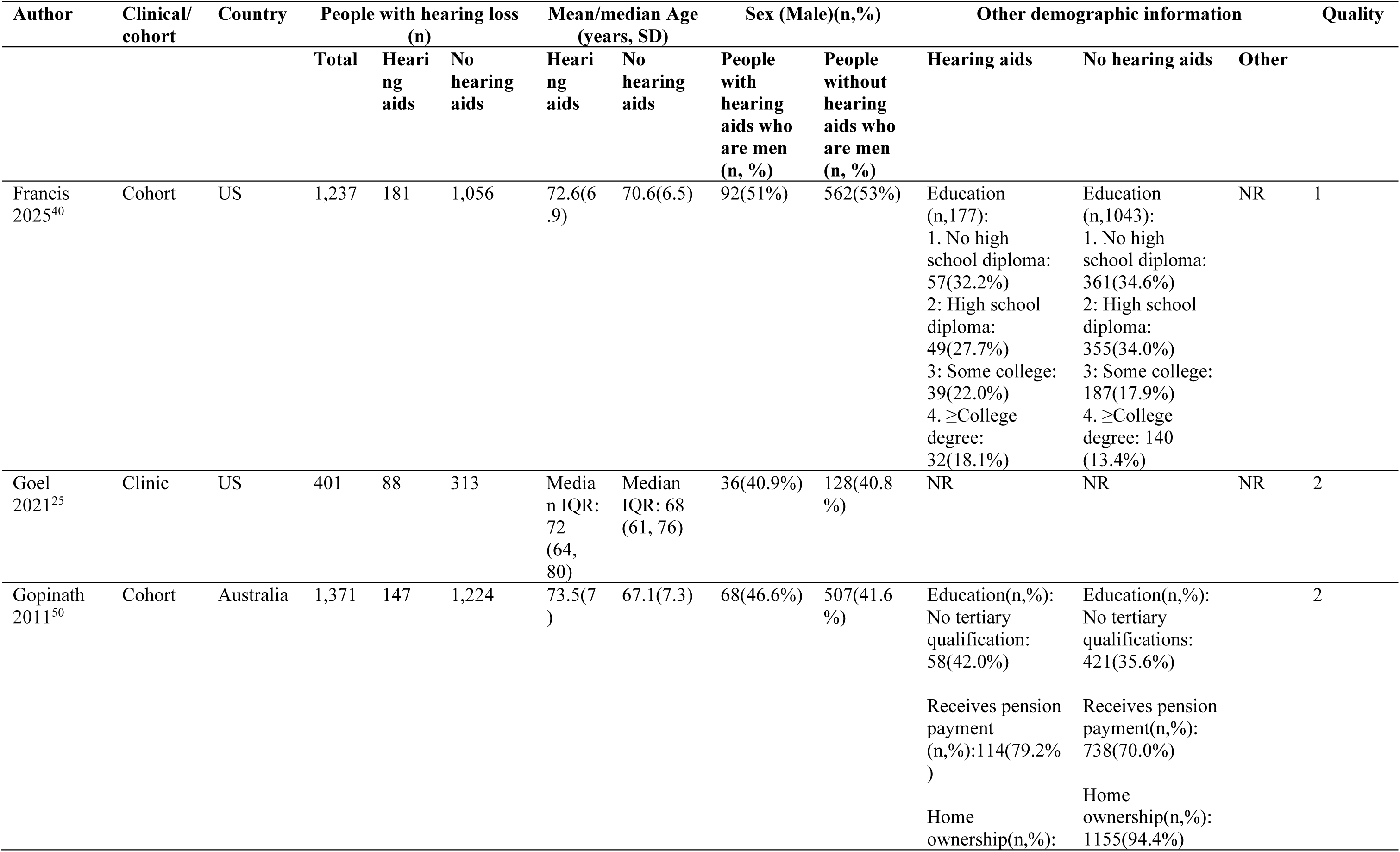
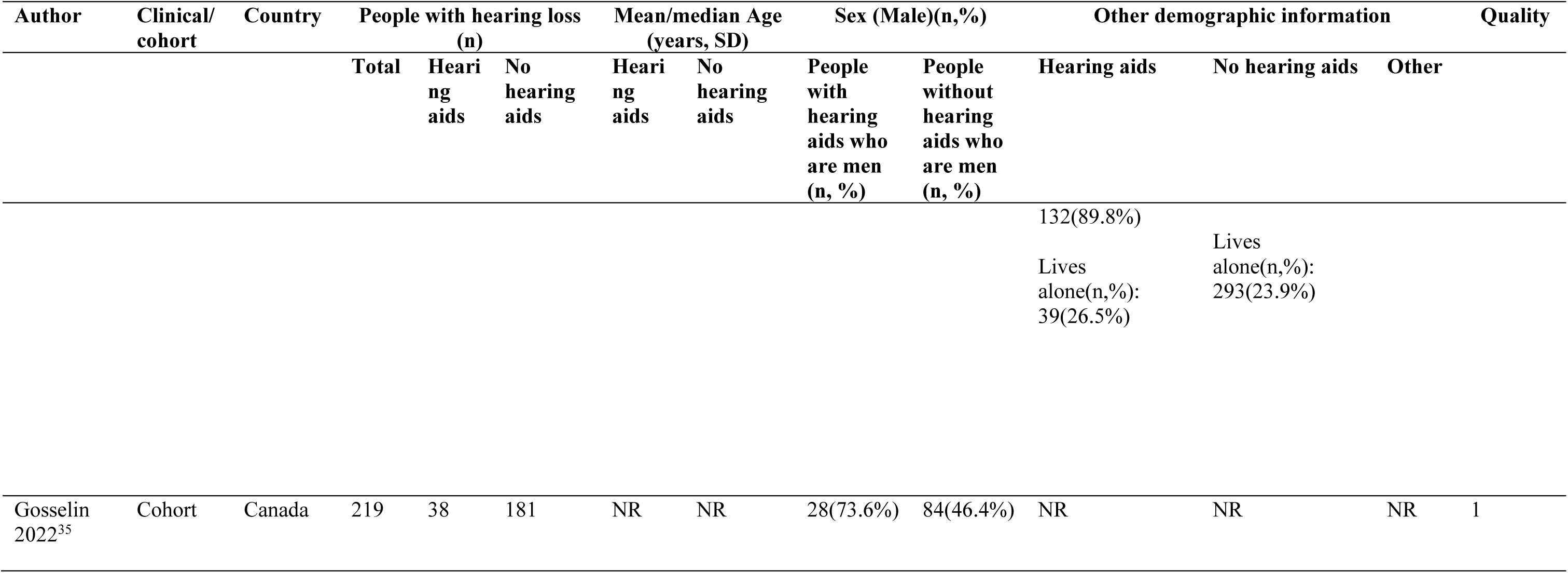
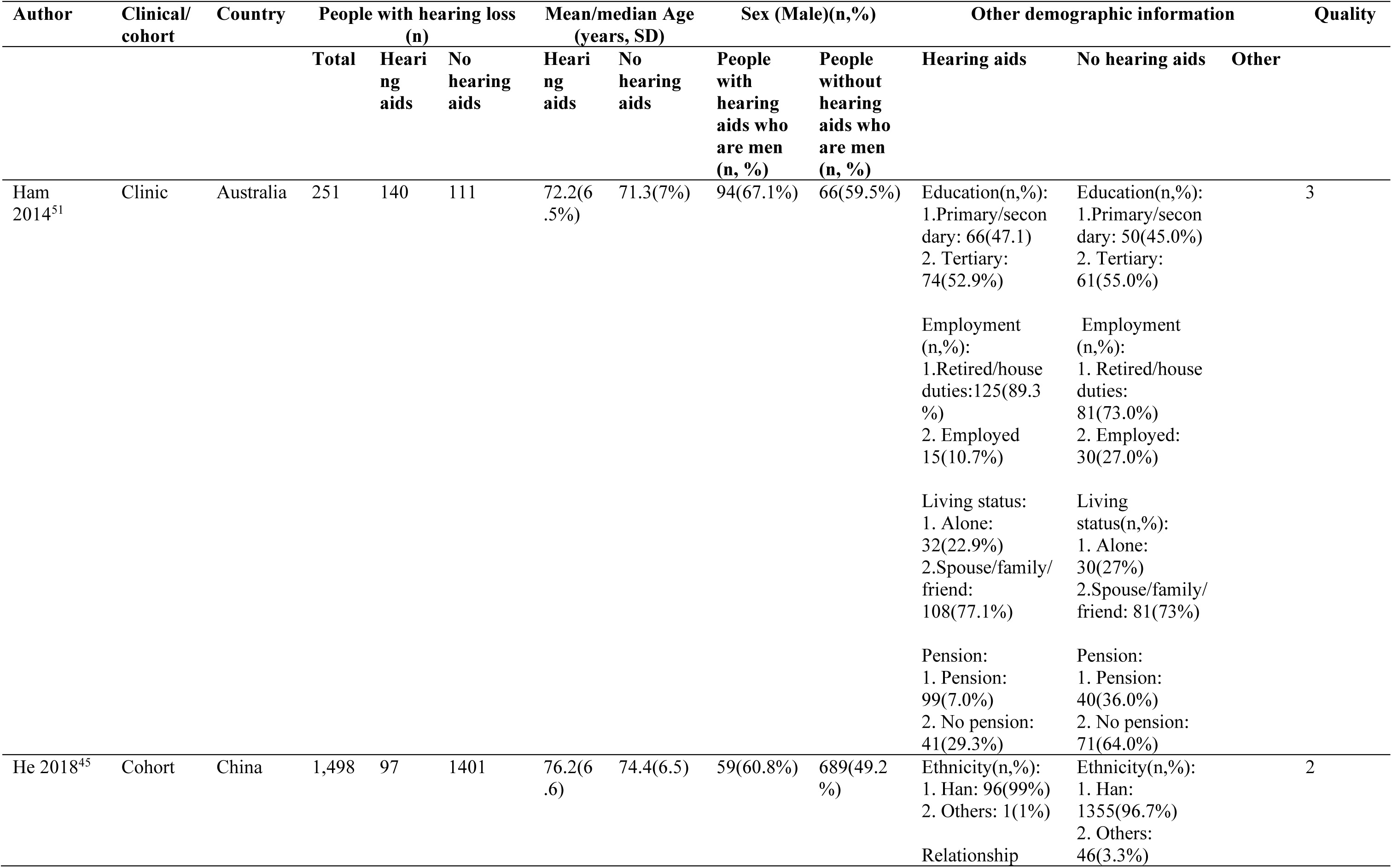
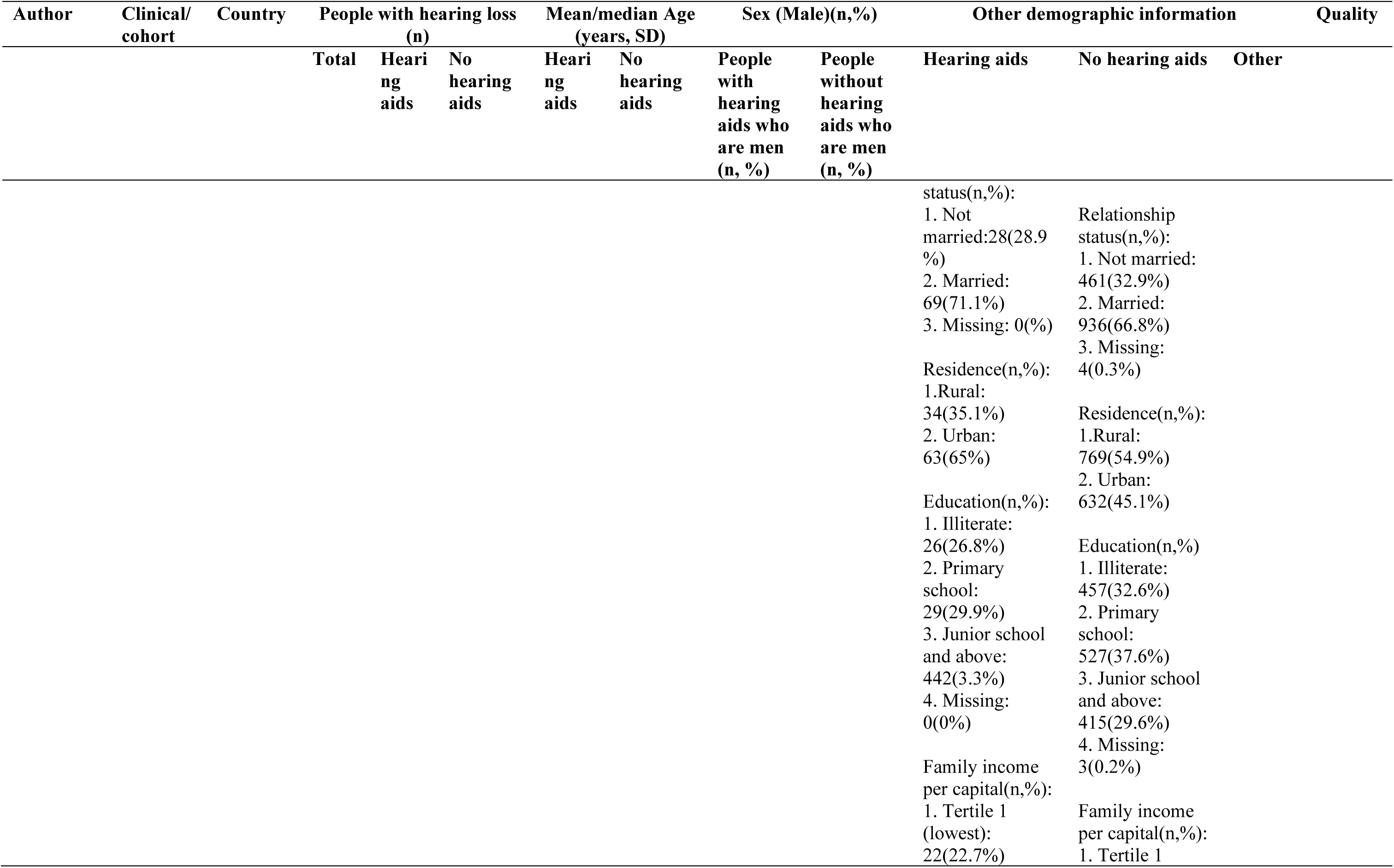
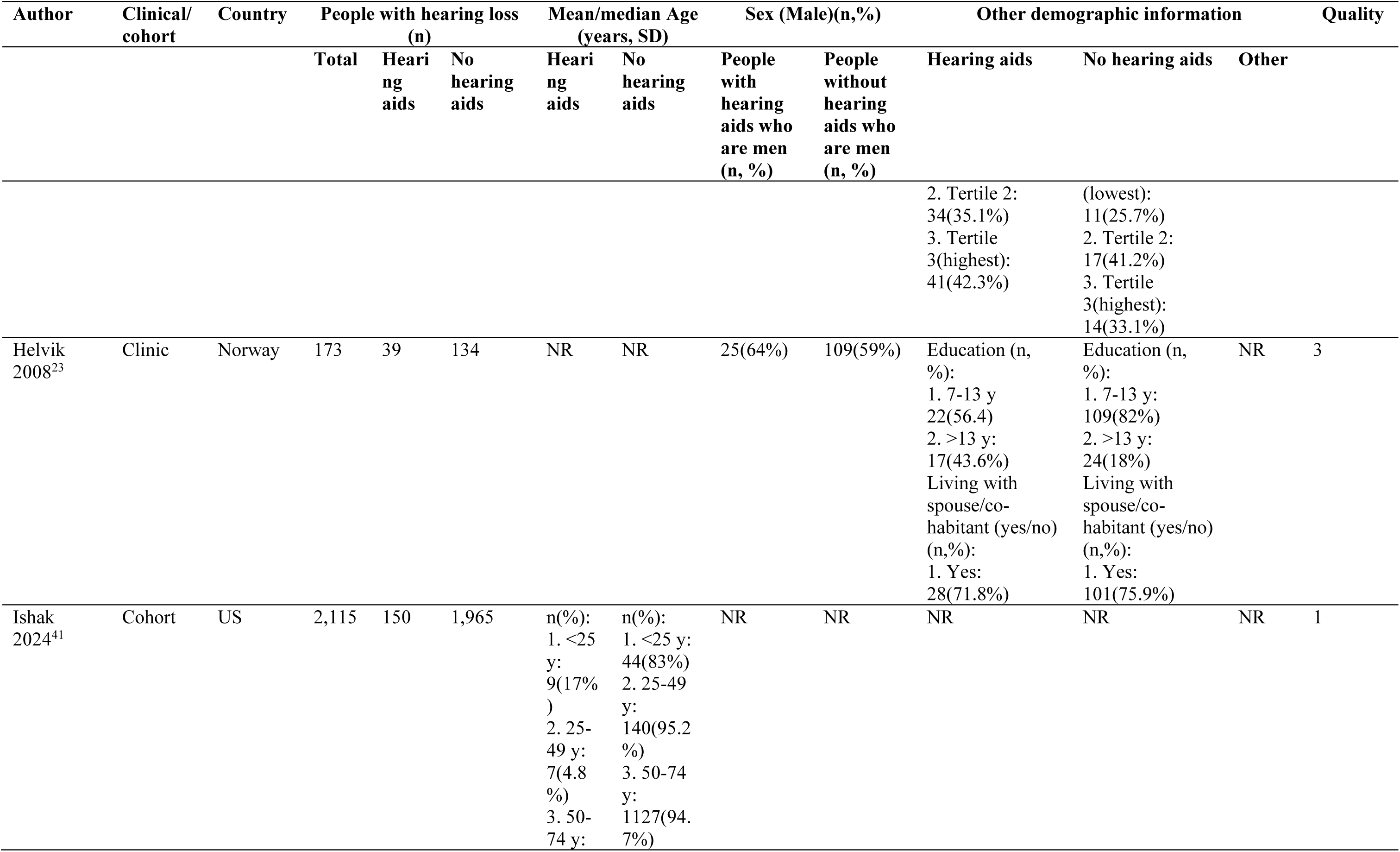
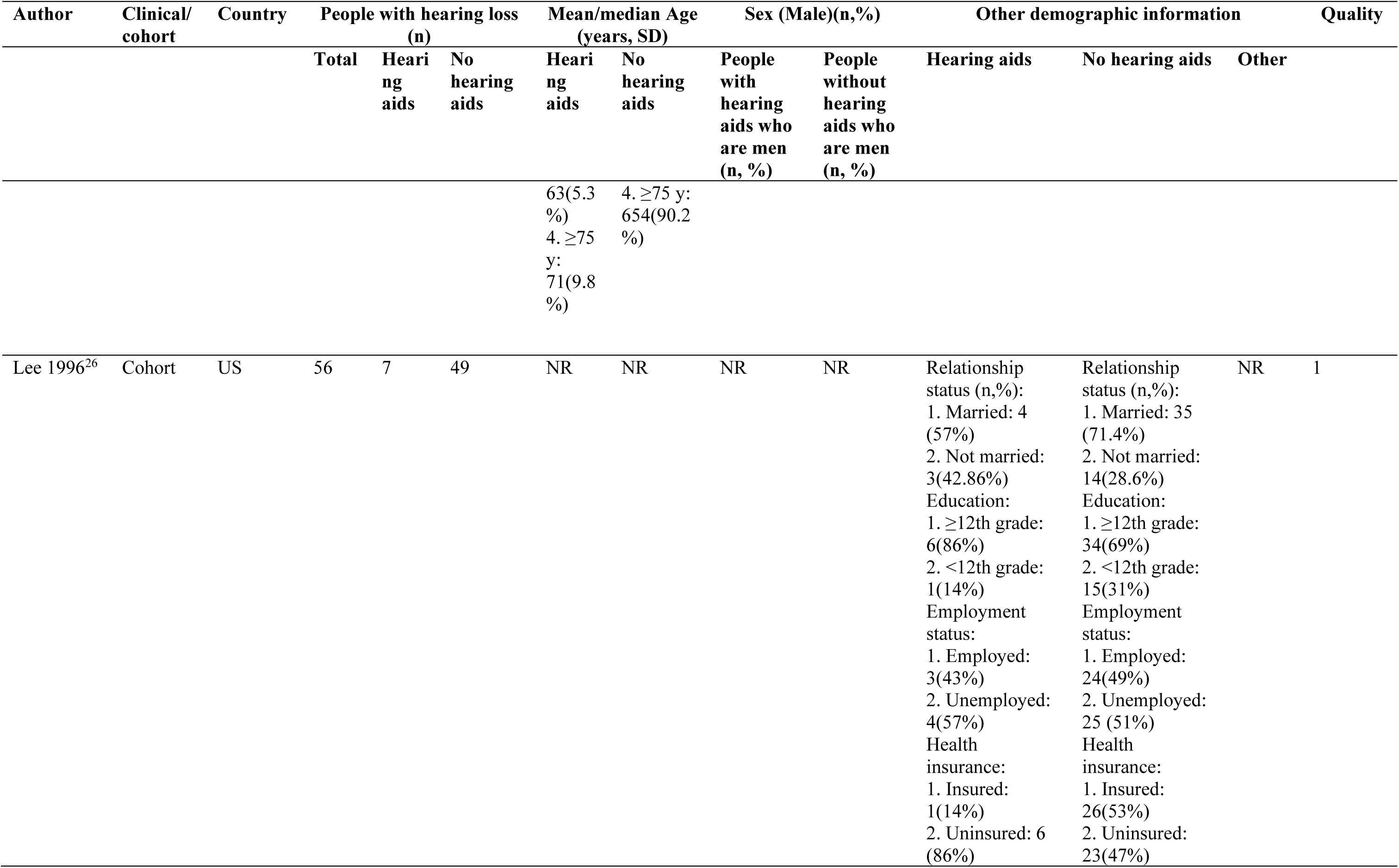
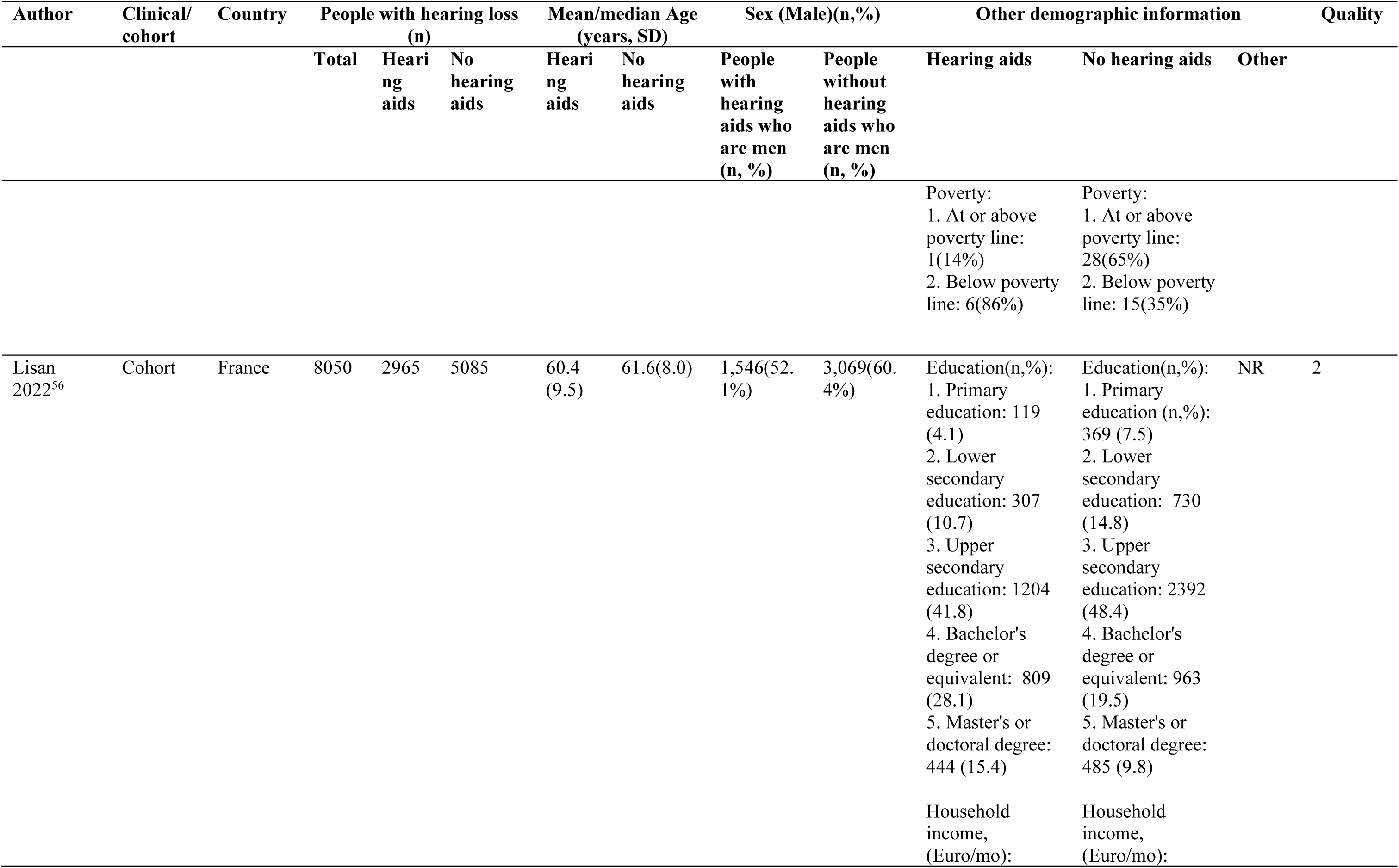
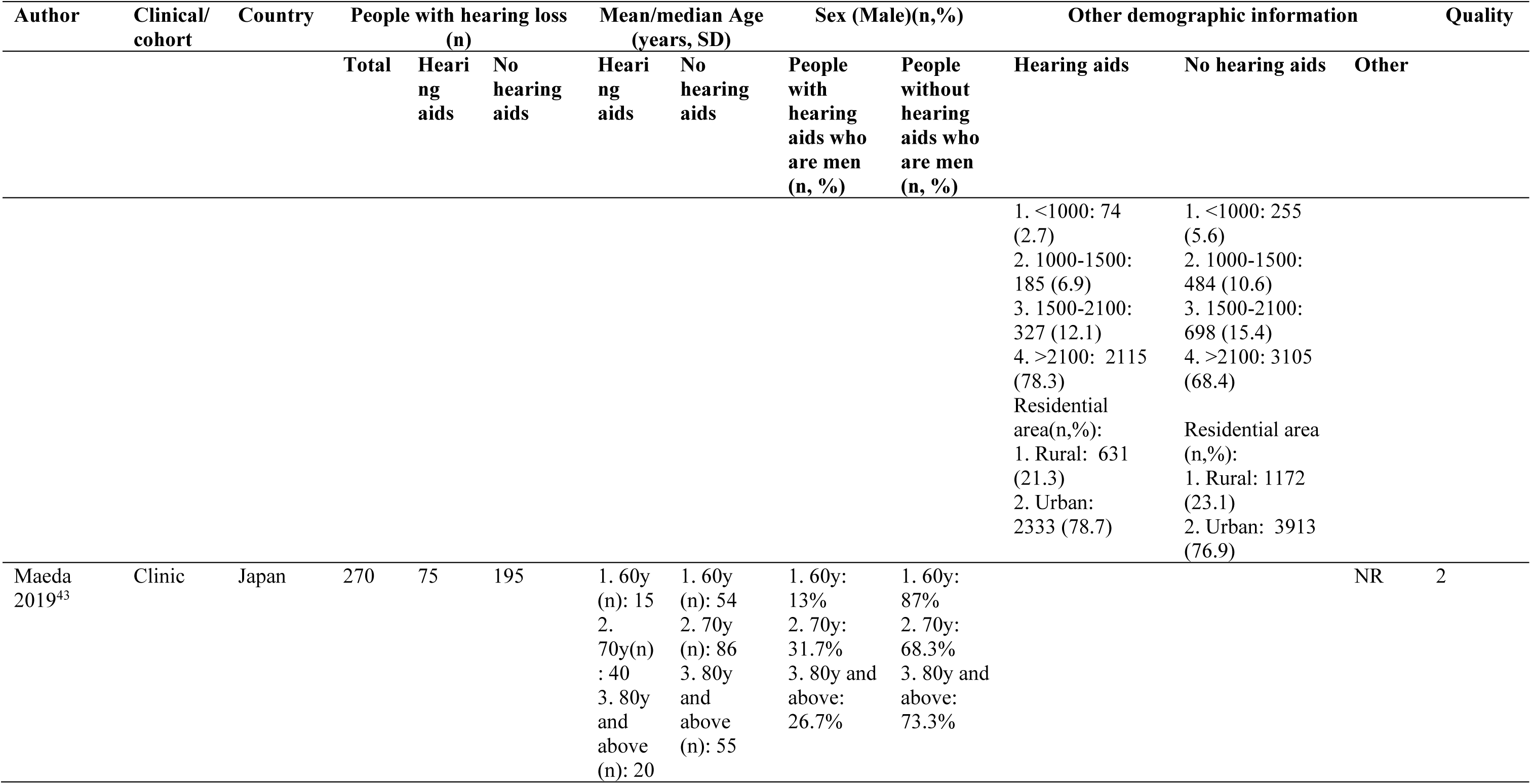
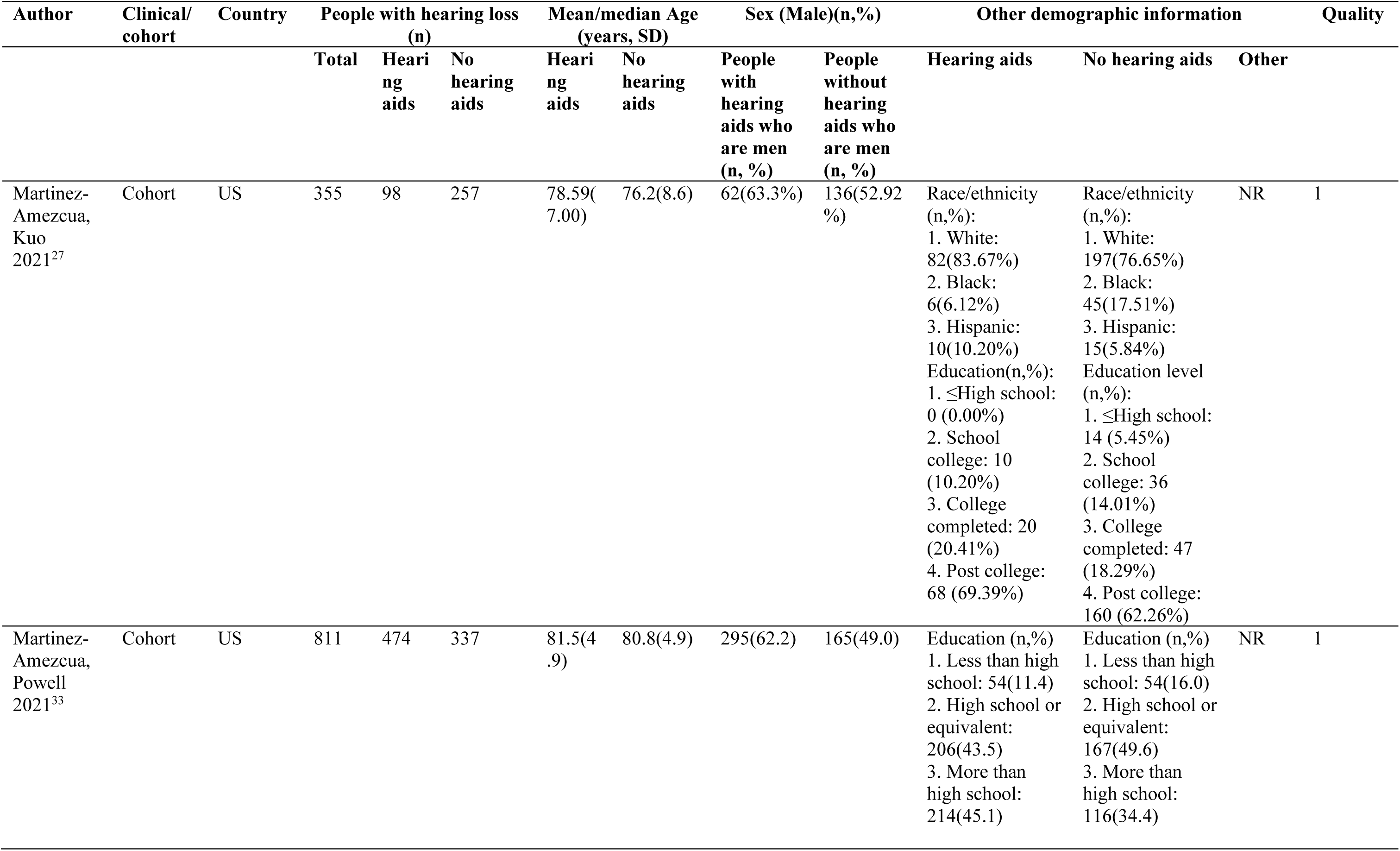
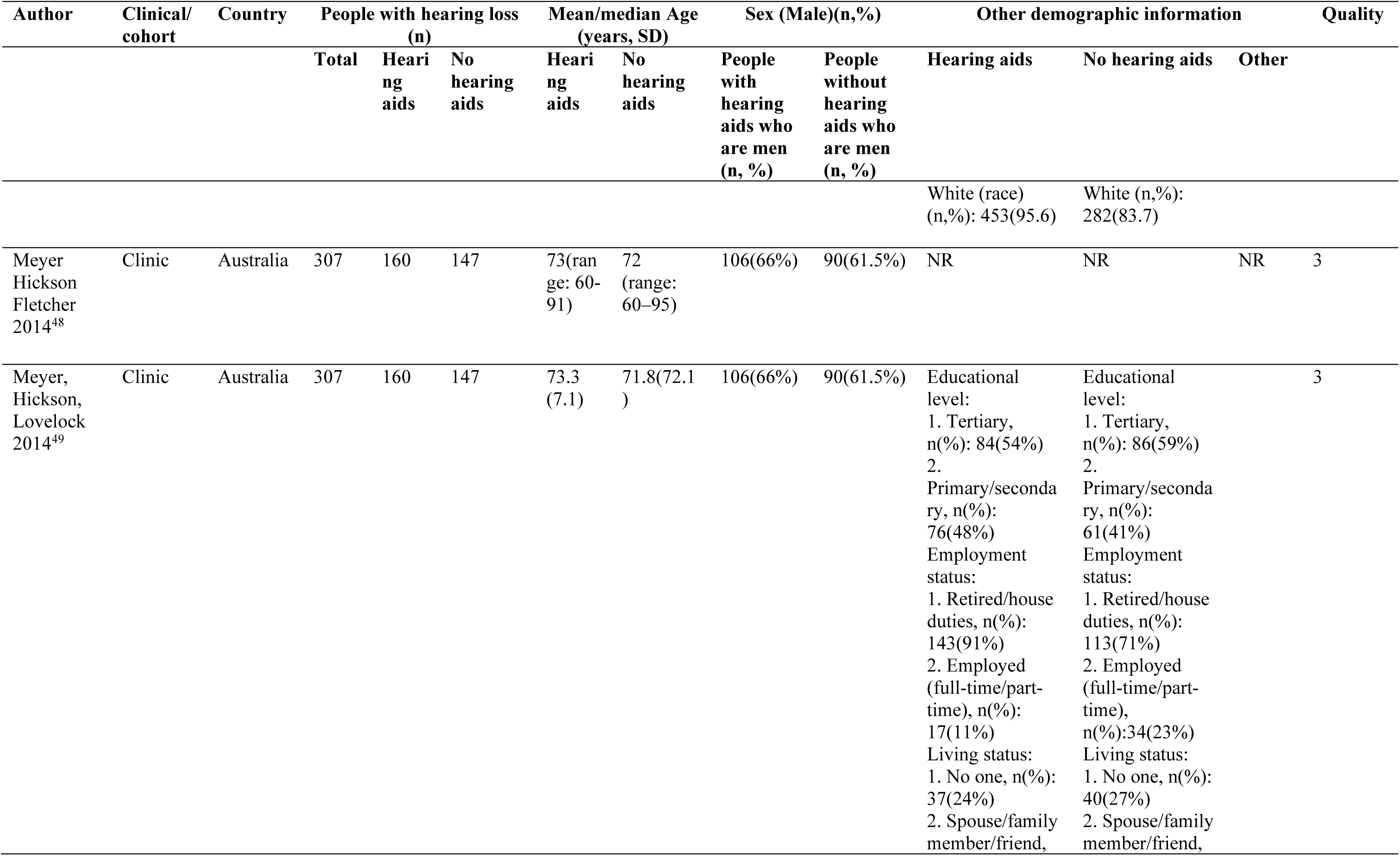
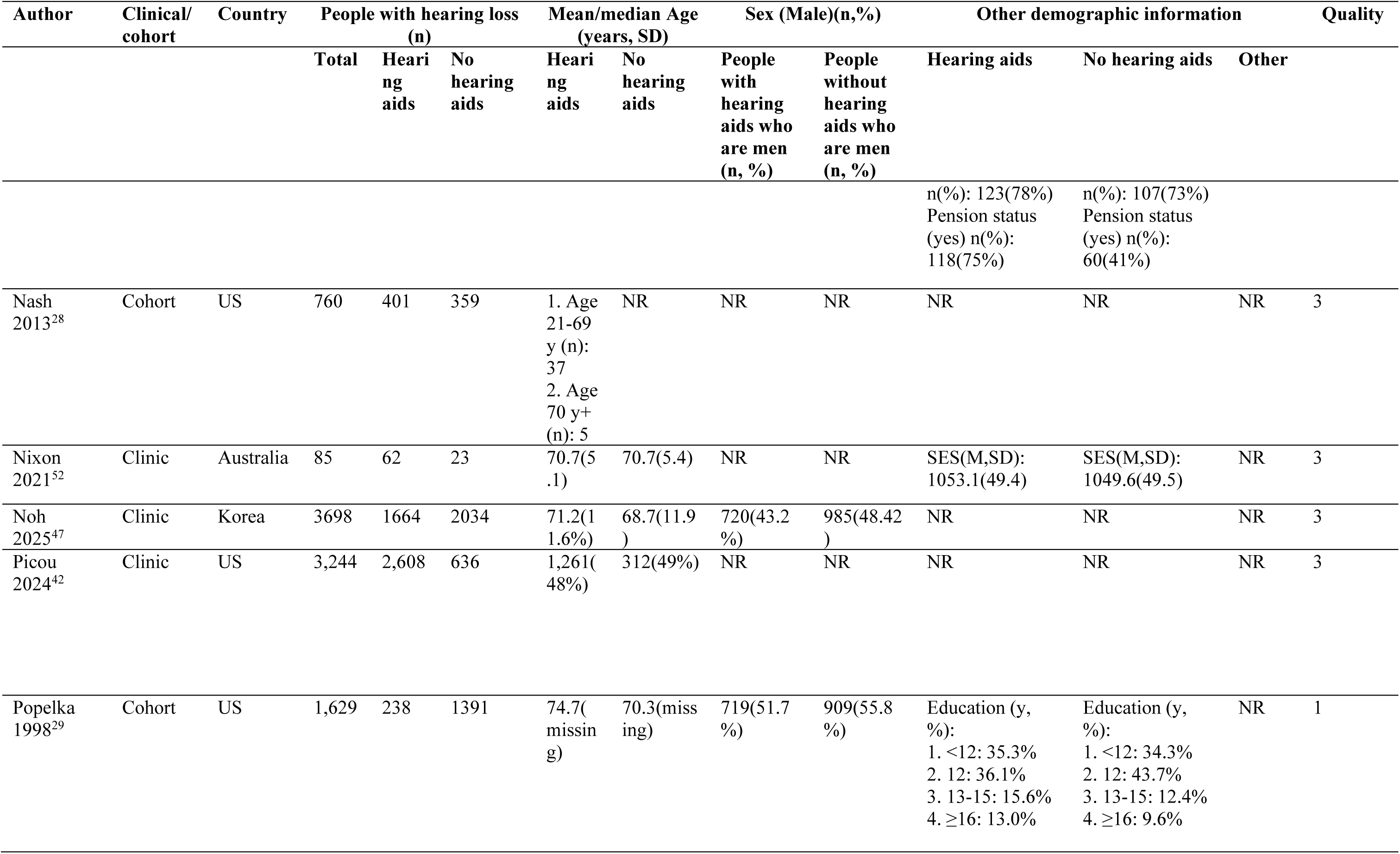
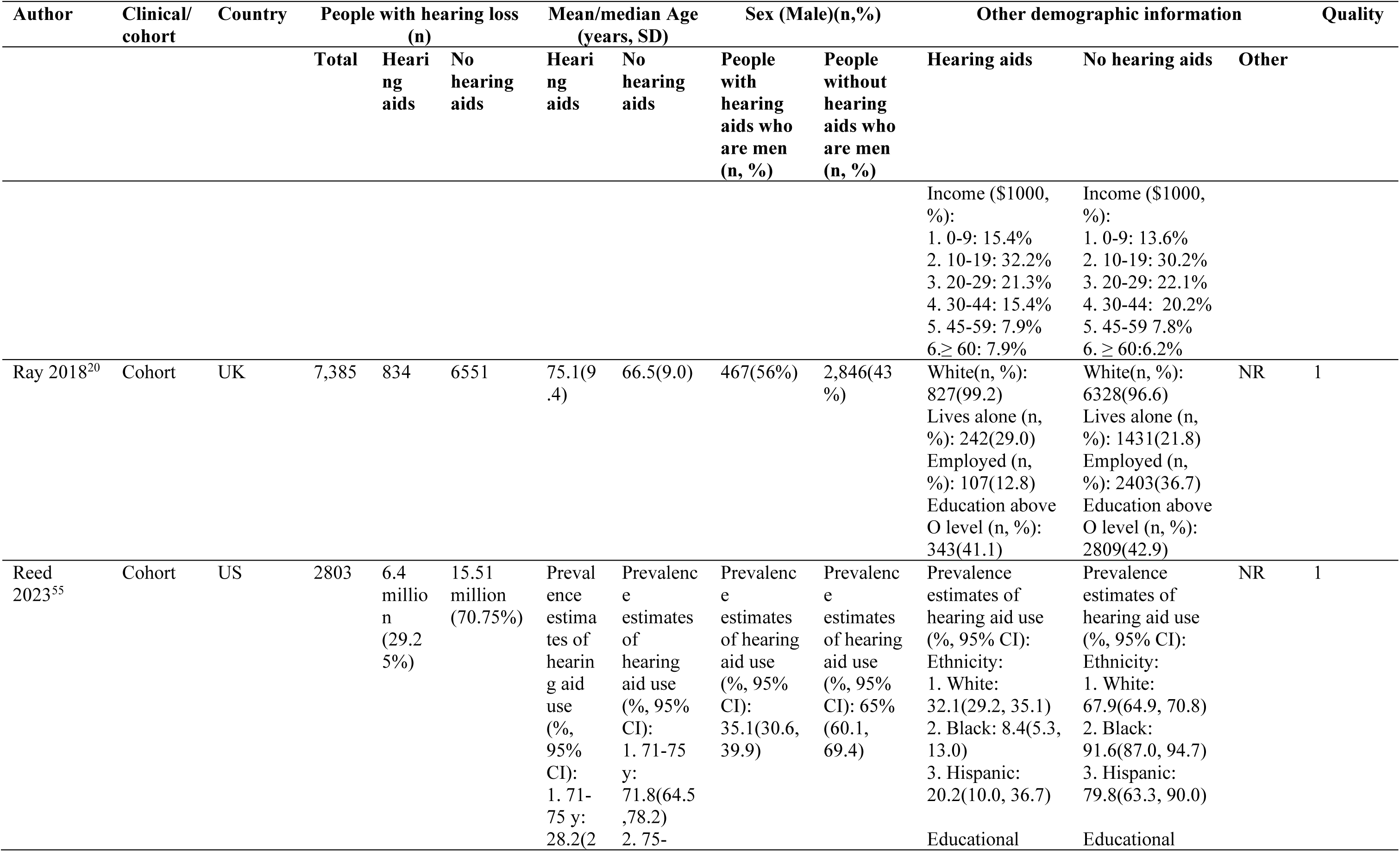
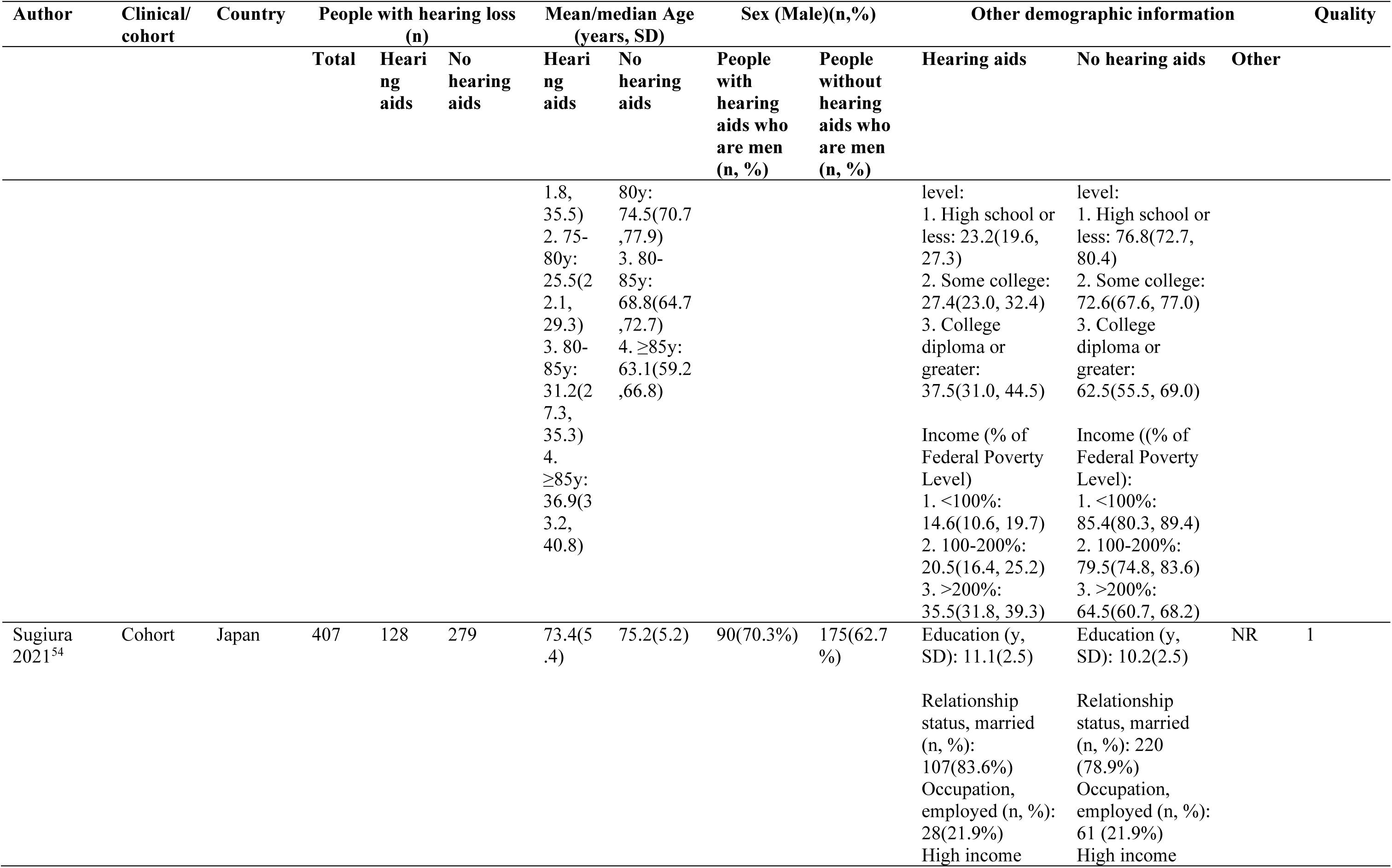
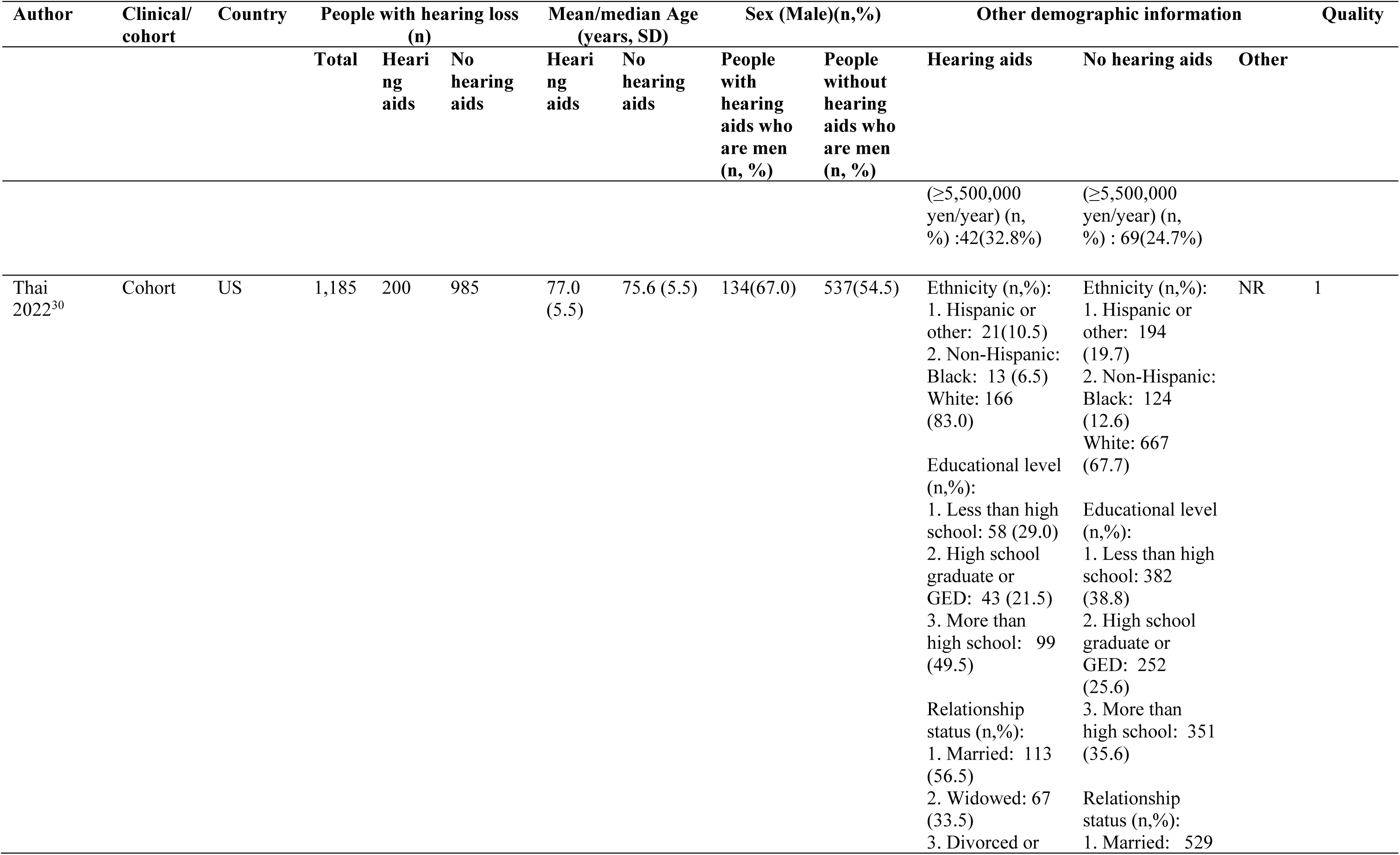
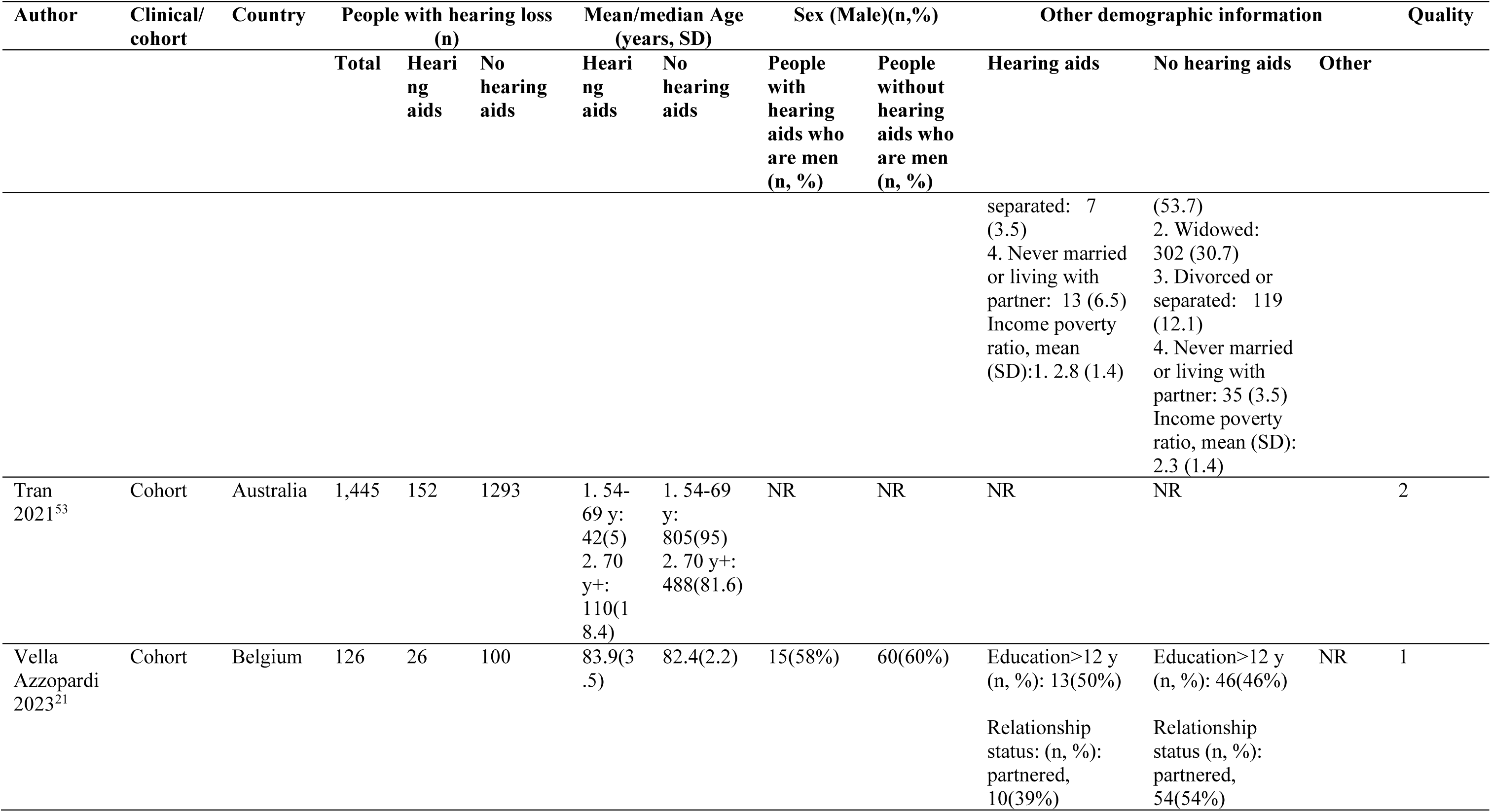
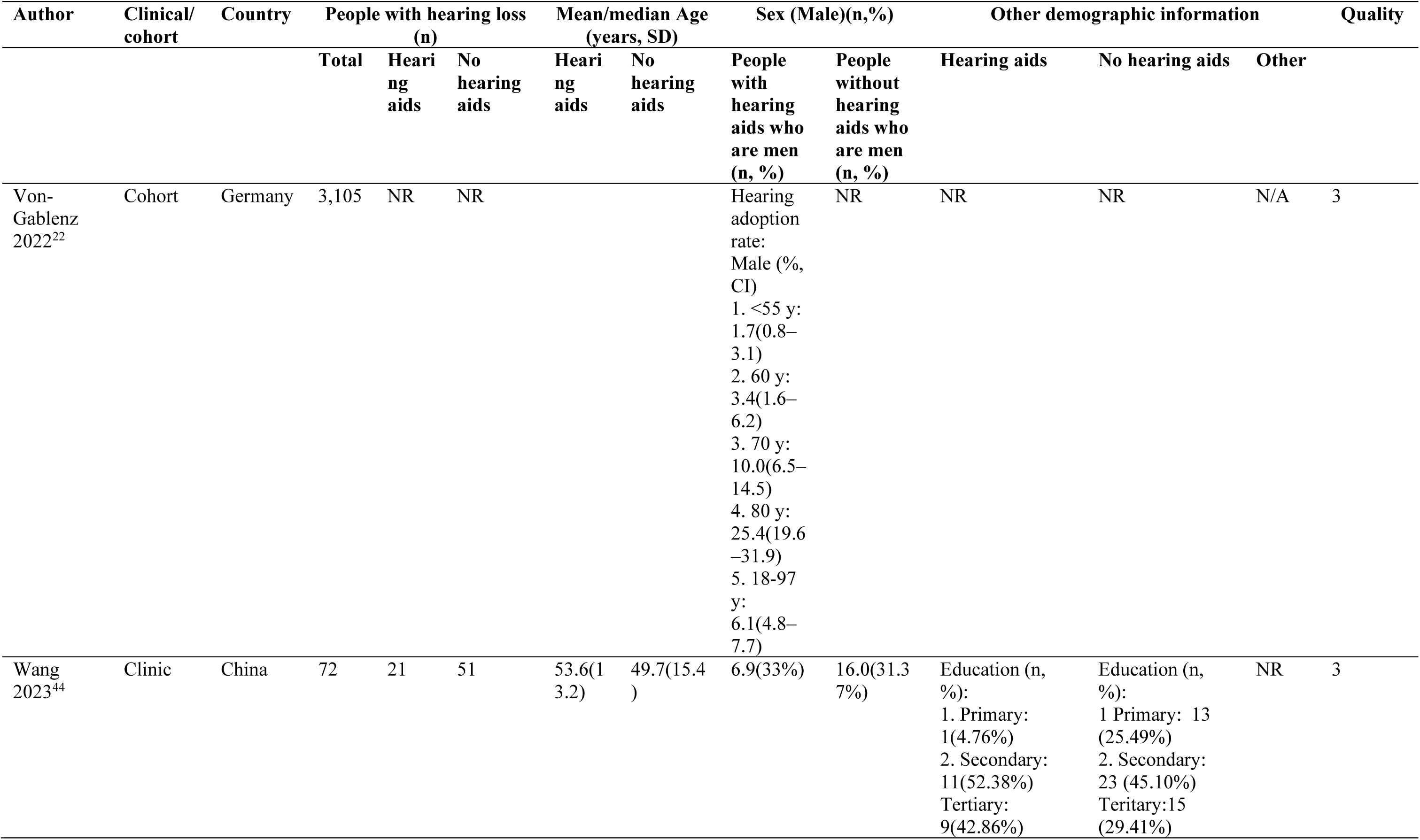
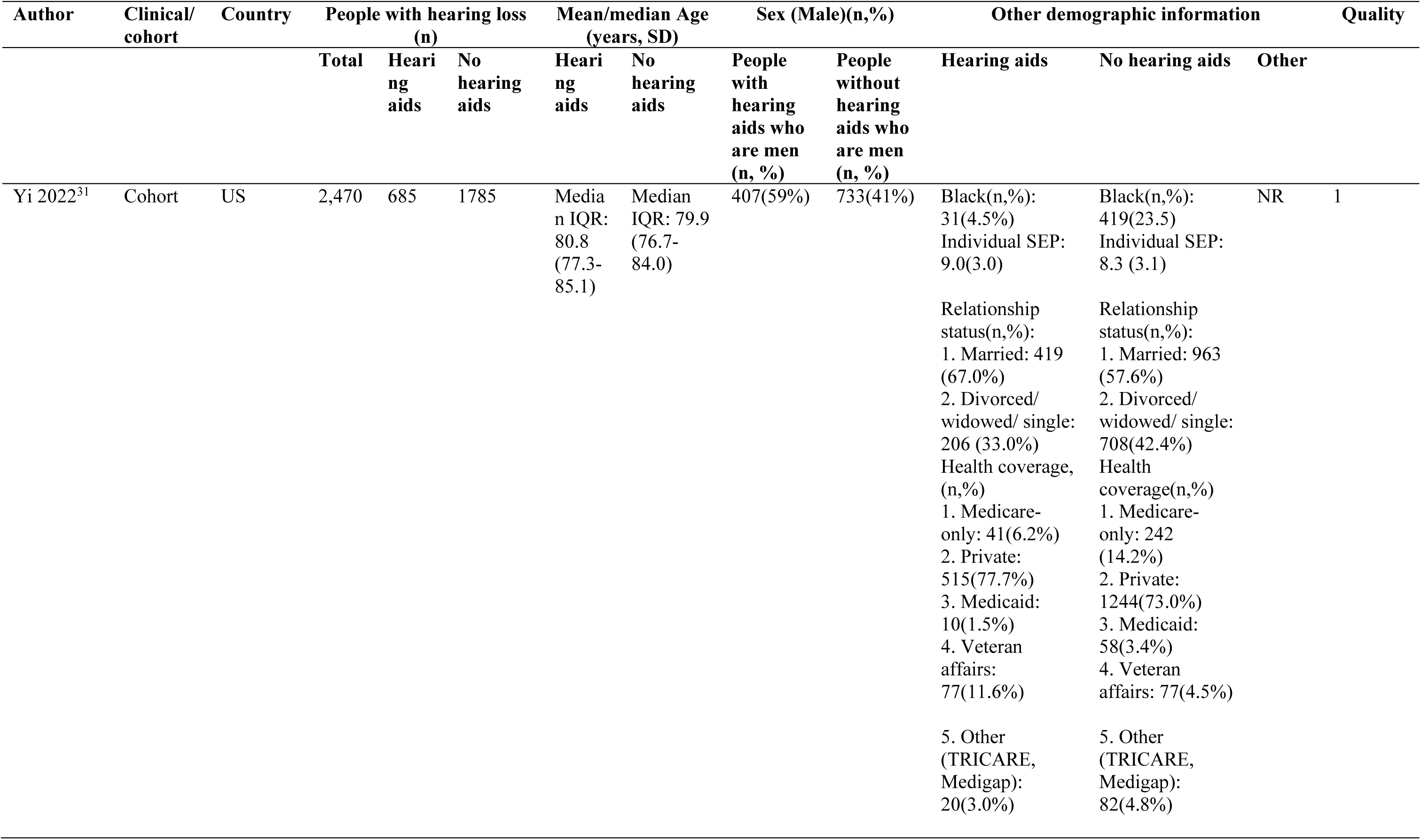
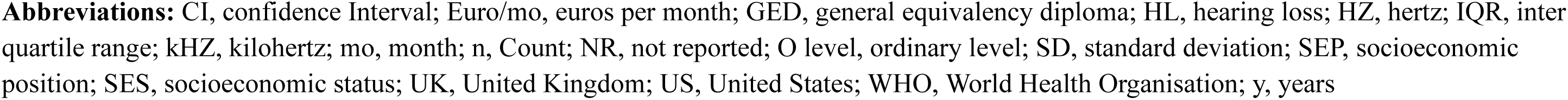
Characteristics of included studies.

### Study characteristics

Of the included studies, six were from Europe,^18–23^ 19 North America,^24–42^ five Asia,^43–47^ and six Oceania (Table 1).^48–53^ None were from low- and middle-income countries. 29 included studies reported on sex,^20–25,27,29–33,35–40,43–45,47–51,54–56^ 32 on age,^20–22,24,25,27–33,36–45,47–56^ 20 on education,^20,21,23,24,26,27,29,30,32–34,36,40,44,45,49–51,54,56^ 10 on ethnicity or race,^20,27,31–33,36–38,45,55^ nine on relationship status (marriage, partnership, or in a relationship),^21,24,26,30,31,36,37,45,54^ six on living with someone or alone,^20,23, 49–51^ and four on employment status.^20,49,51,54^ Other sociodemographic factors reported included socioeconomic status,^18,31,36,52^ rural vs. urban areas,^36,45,56^ insurance coverage (yes/no, types of insurance),^26,31^ pension status,^49,50^ and home ownership.^50^

### Study quality

Eleven studies had an overall score of three,^22,23,28,37,42,44,47–49,51,52^ eight scored two,^24,25,32,37,42,43,45,47,50,53,56^ and 12 scored one^18,20,21,26,27,29–31,33,35,46,55^ out of five. 18/36 studies scored zero on “representativeness”,^18,20,21,24,26,27,29–31,33–36,38,40,41,54,56^ with 11 studies scoring two as participants were recruited from hearing clinics.^22,23,28,37,42,44,47–49,51,52^ Seven cohorts investigating hearing loss scored one for being somewhat representative.^25,28,32,39,45,50,53^ All studies scored one for the ascertainment of exposure items as we were looking for sociodemographic details, and there is no validated tool (Table 1).

Table 1 describes 36 studies from cohorts and eight from clinics, detailing demographic information of 300,946 individuals who do and do not access hearing aids.

### Sex

Of the 29 studies that included sex (n, 293,584),^20–22,24,25,27,29–33,35–40,43–51,55,56^ eight studies were from clinic,^23,25,43,44,47–49,51^ and 21 cohort studies. ^20–22,24,27,29–33,35–40,45,46,50,55,56^ We meta-analysed 24 studies.^22,34,43,49,51^ Two other studies used the same cohort as Meyer, Hickson, Fletcher et al.,^49,51^ and others did not report sufficient information. ^22,34,43^ Men compared to women with hearing loss were more likely to obtain hearing aids, but there was high heterogeneity (n, 287,964; RR, 1.09[95% CI 1.02-1.17]; I^2^, 98.7%) (Figure 2).

**Figure 2.**
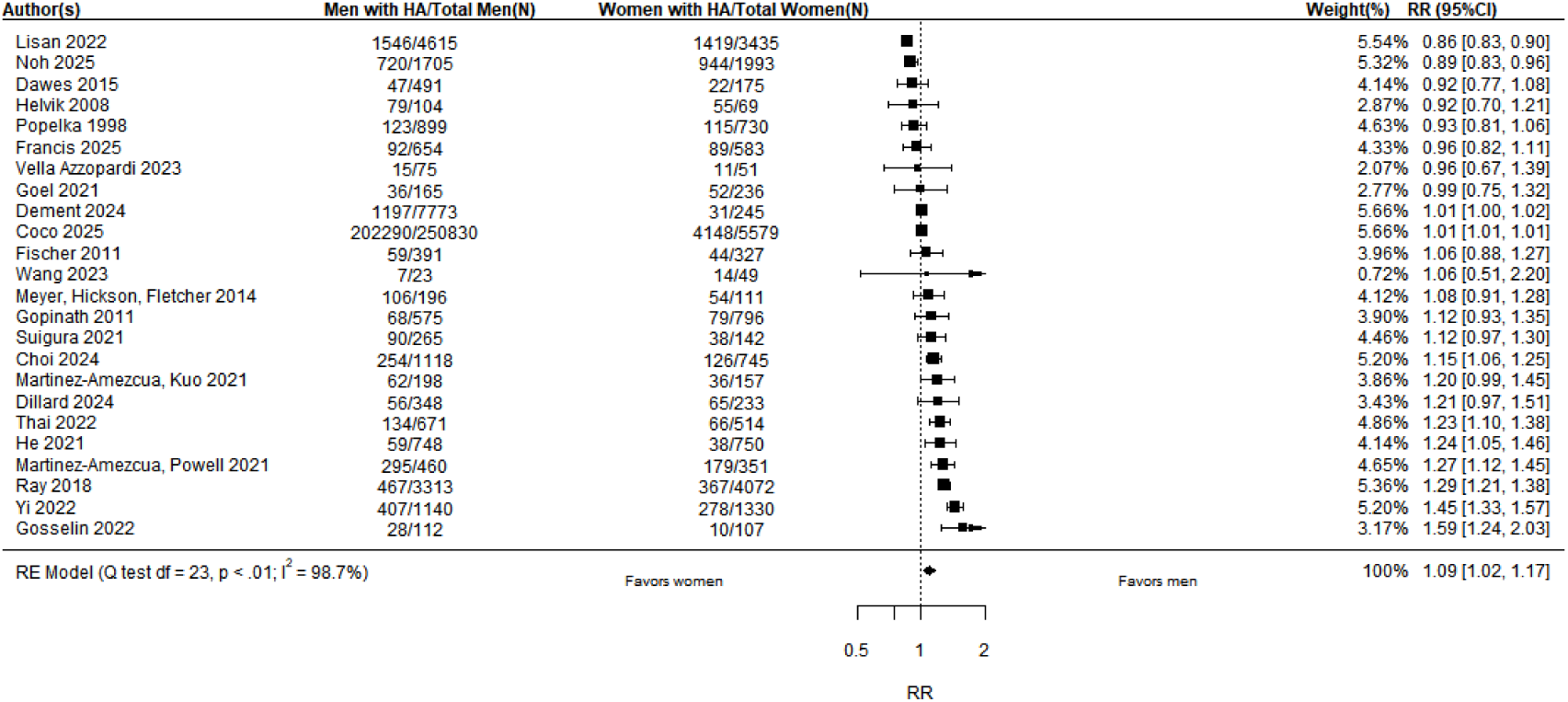
Relative risk of getting hearing aids in men and women, all with hearing loss.

### Ethnicity or race

Although 10 cohort studies reported ethnicity or race (n, 280,446),^20,27,31–33,36–38,45,55^ categories were narrow and non-specific. We meta-analysed seven studies comparing hearing aid access in those who are White versus other ethnicities or race. Two studies could not be categorized like this,^31,45^ and one reported zero people with hearing aids in the non-White category.^32^ White people were more likely to access hearing aids than other ethnicities or races but with high heterogeneity (n, 274,860; RR, 1.26[1.07-1.55]; I^2^, 99.9%) (Figure 3). A sensitivity analysis of studies with White vs. minorities’ ethnicity data only, found RR, 1.12[0.94, 1.33] (n, 8,570; I^2^, 95.2%) (eFigure 1).^20,30^

**Figure 3.**
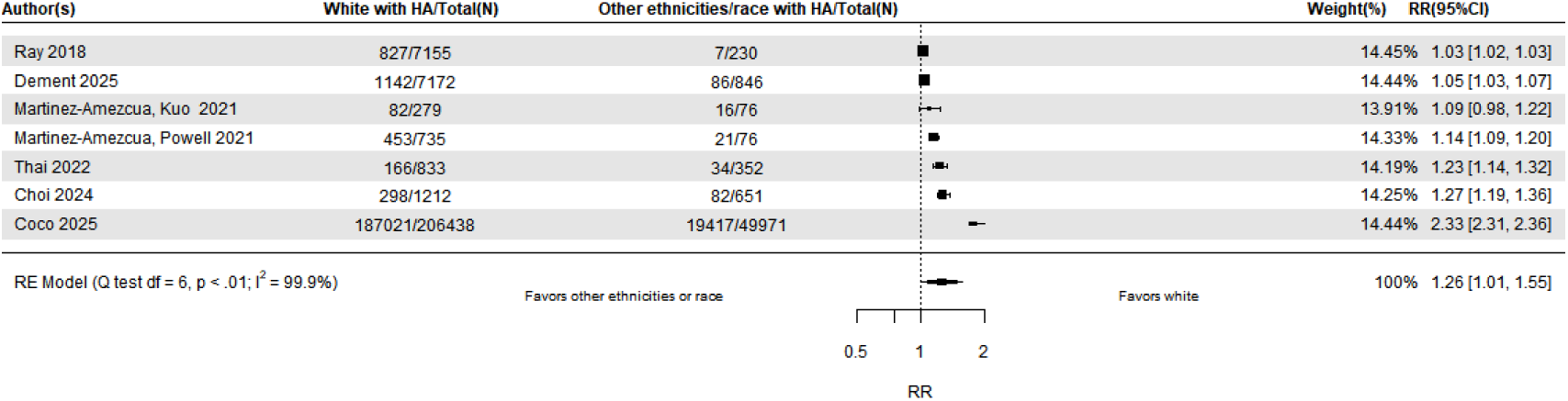
Relative risk of getting hearing aids in White individual and people from other ethnicities, all with hearing loss.

### Education

Twenty studies (n, 18,720) reported education, 16 were from cohorts,^20,21,24,26,27,29,30,32,33,36,40,45,50,54–56^ and four from clinics.^23,44,49,51^ We combined the results from 12 studies to compare individuals with ≥12 years to those with <12 years, finding that people with post-secondary education were more likely to access hearing aids than those without (n, 5,970; RR, 1.17[1.04, 1.32]; I^2^, 79.4%) (Figure 4).^21,24,26,27,29,30,32,36,40,44,49,56^ Seven studies did not report education using the 12-year cut-off points,^20,23,34,45,50,54,55^ Ham et al. analysed the same cohort as Meyer Hickson Lovelock et al.,^51^ and one study was excluded from the meta-analysis as none with hearing aids had less than 12 years of education.^27^

**Figure 4.**
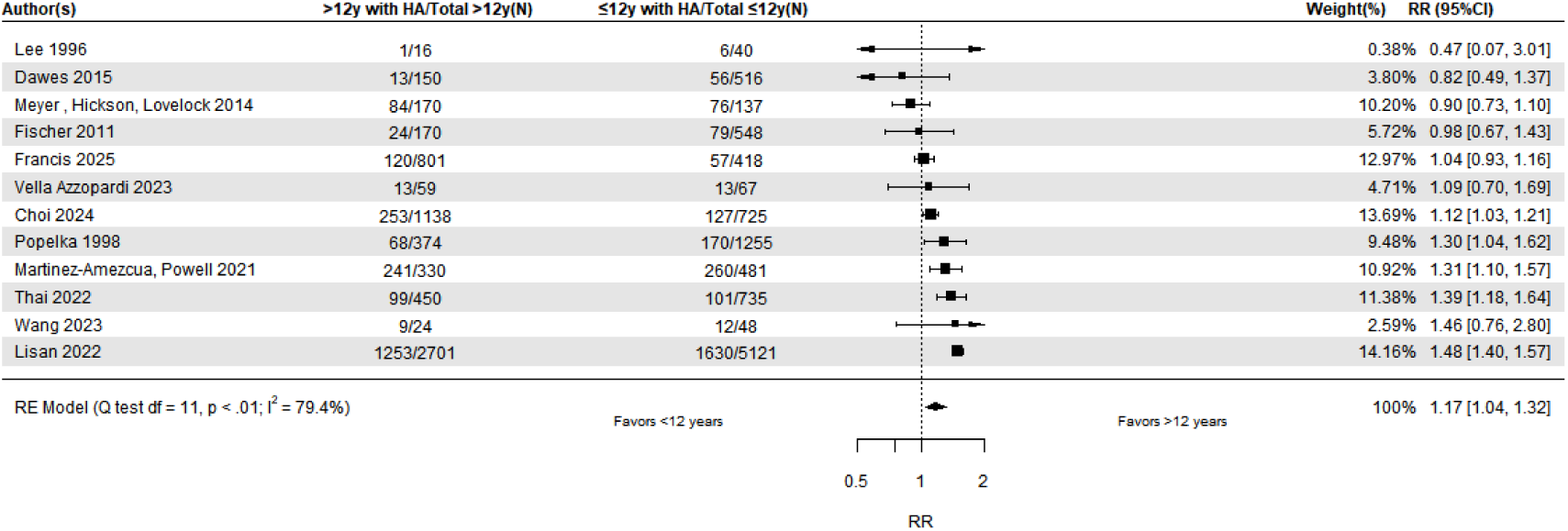
Relative risk of people getting hearing aids with ≥12 and <12 years of education, all with hearing loss.

### Pension status

Two studies, both from Australia had information on pension status.^49,50^ We found those receiving pensions were more likely to acquire hearing aids (n, 1,678; RR, 1.50[1.08,2.09]; I^2^, 87.4%) (eFigure 2 Supplemental 1), but this had high heterogeneity.

### Income

Eight cohort studies (n, 262,339) reported income (Table 1),^24,29,32,45,46,55,56^ and we meta-analysed three studies from the US with the same very low household income breakdown.^24,29,32^ We compared hearing aid access in people with hearing loss with income of ≥$45,000/year to those with <$45,000/year. We did not find that income affected access to hearing aids with low heterogeneity (n, 3,013; RR, 1.06[0.87-1.30]; I^2^, 0%), (eFigure 3 Supplemental 1). In the other five studies,^37,45,46,55,56^ more people with higher income acquired hearing aids than those with lower income (Table 1).

### Employment status

We meta-analysed four cohort studies (n, 7,380) with participants who were currently employed vs. not employed and found no difference in hearing aid access (n, 7,380; RR, 0.58[0.34-1.00]; I^2^, 84.2%) (eFigure 4 Supplemental 1), but heterogeneity was high.^20,46,48,51^

### Relationship status

We meta-analysed nine cohort studies by categorizing them as either currently married/in a relationship/partnered, or not (separated, divorced, single, widowed, or never married).^21,24,26,30,31,36,37,45,46^ We found no difference (n, 263,528; RR, 0.96[0.87-1.16]; I^2^, 70.0%) (eFigure 5 Supplemental 1).

### Living alone or with others

Six studies reported whether participants lived alone or with others.^20,23,31,49–51^ One cohort was in three papers,^48,49,51^ and we included only one.^49^ There was no difference in meta-analysis of four studies (n, 8,756; RR, 0.82[0.57-1.18]; I^2^, 97.7%) (eFigure 6 Supplemental 1).^20,23,49,50^

### Rural vs. urban areas

We meta-analysed two studies comparing participants living in rural vs. urban areas, and found no difference (n, 9,548; RR, 1.20[0.86-1.68]; I^2^, 94.4%) (eFigure 7 Supplemental 1).^45,56^

## Discussion

This is, to our knowledge, the first systematic review meta-analysing the sociodemographic characteristics of people who do and do not get hearing aids and equality in hearing aid access. All studies were observational with cross-sectional information about acquiring hearing aids. We found that men were 9% more likely to access hearing aids than women, and White people were 26% more likely than other ethnicities or races. People with more than 12 years’ education were 17% more likely to get hearing aids, and pension recipients were 50% more likely, compared with those with less education and non-recipients, though these findings may differ between populations and may limit external validity. It also suggests that these demographic barriers can be overcome in some circumstances. We did not find an association between hearing aid access and household income, current employment, current relationship status, living arrangement and rural vs. urban areas.

### Interpretation of findings

Jenstad et al.^13^ reported narratively that age and sex were associated with hearing aid uptake, and as the degree of hearing loss increased, women but not men were more likely to acquire a hearing aid. Another systematic review reported mixed findings for sex,^11^ and a literature review reported that sex did not affect hearing aid uptake, and findings on the impact of education level were conflicting.^14^

In this review, we found that men were more likely to access hearing aids than women, in a meta-analysis with people from North America, Asia and Europe. A previous systematic review found no association between sex and adopting hearing aids in 16 out of 20 studies but three found men were more likely to adopt hearing aids.^11^ Past studies have focused on the usage of hearing aids rather than access to them. Hearing aid access refers to whether someone gets the hearing aid, and adoption includes both the uptake and use of hearing aids. While it is essential that individuals obtain hearing aids to use them, access and usage are distinct, and findings are conflicting. For example, the US National Health Interview Survey reported that 7.1% of adults aged ≥45 used a hearing aid, and this was higher among men than women at all ages.^57^ However, a cross-sectional survey of 8,389 people in Switzerland found that women used hearing aids more regularly and had a longer daily duration of use.^58^

We also found White people were 26% more likely to access hearing aids than other groups, similar to a recent US survey investigating access to primary and preventative care in 50 states.^59,60^ They reported minorities, including Black, Hispanics, and Native Americans, receive worse healthcare than White individuals. This is similar in the UK where there are ethnic inequalities across a range of healthcare services, with Black communities having especially poor access and outcomes.^61,62^ Reasons for poorer access for minorities could include a lack of appropriate interpreting services, delays and avoidance of seeking help due to fear or racism.^62^

Our finding that individuals with post-secondary education were more likely to get hearing aids than those without aligns with other studies indicating that education is strongly associated with health behaviours.^63,64^ Education is both a driver of opportunity and a contributor to inequality.^65^ so people who are more educated generally age more healthily,^64^ Individuals with less educational attainment are also less likely to get a recent hearing test, ^66^ which may further affect whether individuals with hearing loss obtain a hearing aid.

Those who received pensions in Australia (where pension age is 67) were more likely to get hearing aids than those who did not, in line with a past systematic review that receiving pensions was linked to increased hearing aid uptake.^11,67^ It could be that more pension recipients are getting hearing aids because they are older, and thus, their hearing impairment is more severe as severity is associated with age.^68^ In Australia, hearing aids can be subsidised. Pension recipients may have greater financial flexibility, enabling them to afford higher-quality, more suitable hearing aids, possibly increasing the likelihood of acquiring one. However, we would need more information; for example, whether these pensions are private or public, and whether people who are not receiving pensions are still working.

### Strengths and limitations

Our findings, from various countries, provide the most up-to-date evidence, and our numbers are large. The systematic nature of this review allowed us to give a comprehensive evaluation of the sociodemographic characteristics of hearing aid access. However, most meta-analyses had high heterogeneity, suggesting differences between settings.

Like education, income was not recorded comprehensively in the included studies. The highest cut-off point for household income was US$45,000/annum, but this is well below the national average income in the US (US$80,610 in 2023).^69^ For pension status, which may be linked to income, the number of studies and people included in our analysis was small, which also limits generalizability. While we did not find an overall impact of very low household income on hearing aid access, our findings are highly heterogeneous, possibly depending on the cost of hearing aids. For example, Medicare and most insurance plans do not cover the cost of hearing aids in the U.S.^70^ In Japan, most people pay out of pocket, with public insurance partially covering the cost for children and those with disabling hearing loss.^71^ In Germany, only individuals earning less than 60,000 euros/annum can obtain hearing aids through public coverage.^71^ However, the UK National Health Service provides hearing aids for everyone, with only a minority choosing to purchase premium versions privately.^62^

Most of the included studies did not record specific ethnicities or racial groups; hence, we categorized them as White versus other ethnicities or racial groups for the meta-analysis. Two of the three studies that categorized participants as White or Black,^20,31,32^ had almost exclusively White participants (e.g., 99%).^20,32^ This limits the generalizability and representativeness of our findings.

Since all the studies were observational and none controlled for hearing severity, it is possible that men, White participants, those with higher education and pension recipients accessed testing later and had worse hearing, and of confounding by indication. It is unlikely to explain everything. All participants in this review either participated in a cohort study or had access to a hearing aid clinic. Those who are not involved in research or cannot access healthcare services may have greater difficulty in obtaining a hearing aid, and they were not included in this study. Another limitation is that we could not consider the source of heterogeneity because of the few included studies in each meta-analysis.

Some included studies were not in our database searches because observational terms were not in their titles or abstracts. To mitigate this, we screened reference lists from previous systematic reviews and included studies.

### Implications and conclusion

Overall, there is evidence suggesting that women, people with less education, those from minority ethnic or racial groups and individuals not receiving pensions with hearing loss are less likely to access hearing aids after hearing tests. However, studies were observational and are limited by confounding by indication. Due to the high heterogeneity in meta-analyses and few studies of high quality, our findings may not be externally generalizable, and it is hopeful that they suggest that barriers to accessing hearing aids can, in some circumstances, be overcome.

The heterogeneous findings suggest that future qualitative work could provide information about barriers and facilitators to hearing aid access, as sociodemographic status does not consistently explain access.

## Data Availability

All data produced in the present study are available upon reasonable request to the authors

## Author Contributions

Dr. Esther K. Hui had full access to all of the data and takes responsibility for the integrity of the data and the accuracy of the data analysis.

*Concept and design:* Livingston, Mukadam, Hui

*Acquisition, analysis, or interpretation of data:* Livingston, Mukadam, Hui, Wang, Marston, Rossetti

*Drafting of the manuscript:* Hui, Livingston, Mukadam

*Critical review of the manuscript for important intellectual content:* All authors

*Statistical analysis:* Hui, Marston, Livingston, Mukadam

*Administrative, technical, or material support:* Hui, Wang, Rossetti

*Supervision:* Livingston, Mukadam, Hui

## Role of the Funder/Sponsor

The funder had no role in the design and conduct of the study; collection, management, analysis, and interpretation of data; preparation, review, or approval of the manuscript; and decision to submit the manuscript for publication.

Disclaimer: The findings and conclusion of this review are those of authors and does not necessarily reflect the view of the NIHR

## Data Sharing Statement

See Supplement 2

