## Supplemental 1 for "Equality in Hearing Aid Access: A Systematic Review and Meta-analysis"

**Supplemental Online Material**

**List of tables and figures**

eFigure 1. Relative risk of people getting hearing aids in White individuals and people of other ethnicities, all with hearing loss

eFigure 2. Relative risk of getting hearing aids in pension recipients and non-recipients, all with hearing loss

eFigure 3. Relative risk of people getting aids with ≥$45K/year and <$45K/year, all with hearing loss, all with hearing loss

eFigure 4. Relative risk of people getting hearing aids who are currently employed and not employed, all with hearing loss

eFigure 5. Relative risk of people getting hearing aids who are currently in a relationship and not in a relationship, all with hearing loss

eFigure 6. Relative risk of people getting hearing aids living with others and alone, all with hearing loss

eFigure 7. Relative risk of people getting hearing aids living in urban and rural areas, all with hearing loss

eTable 1. MOOSE checklist

eTable 2. Search Strategy

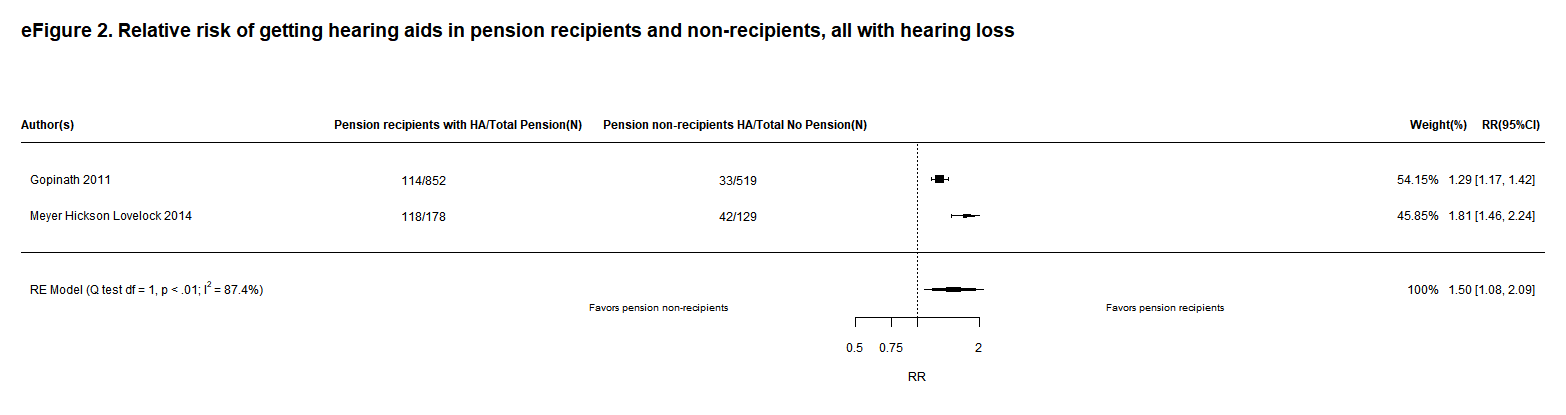

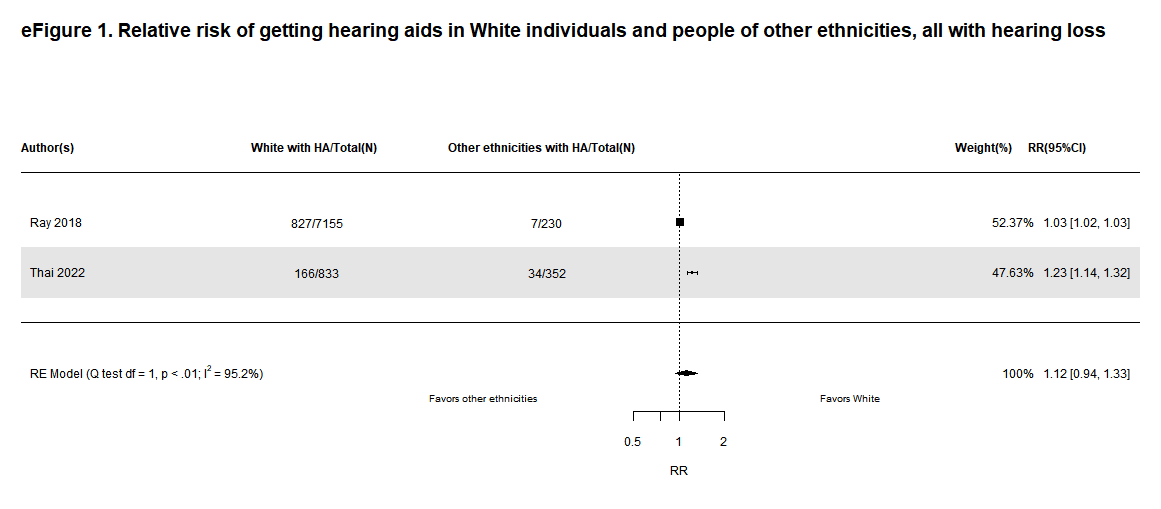

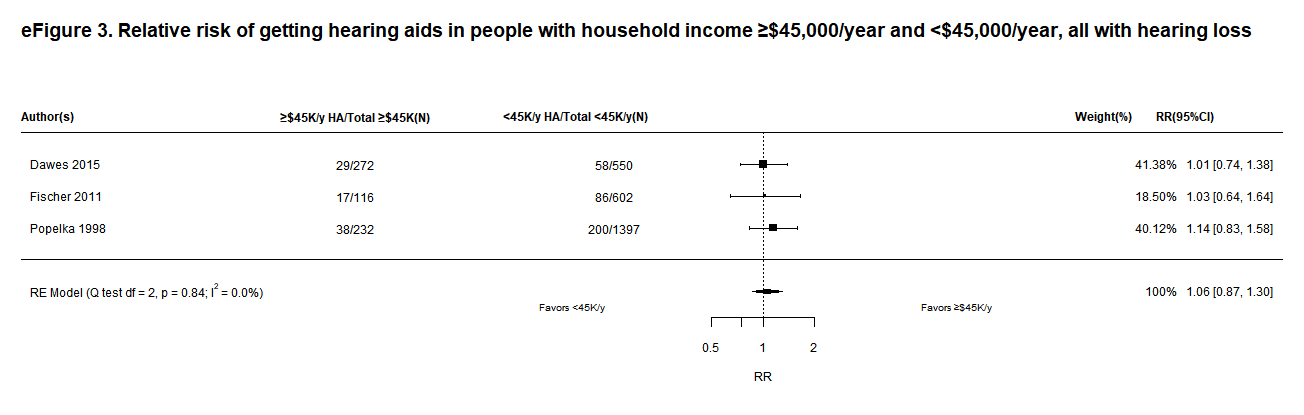

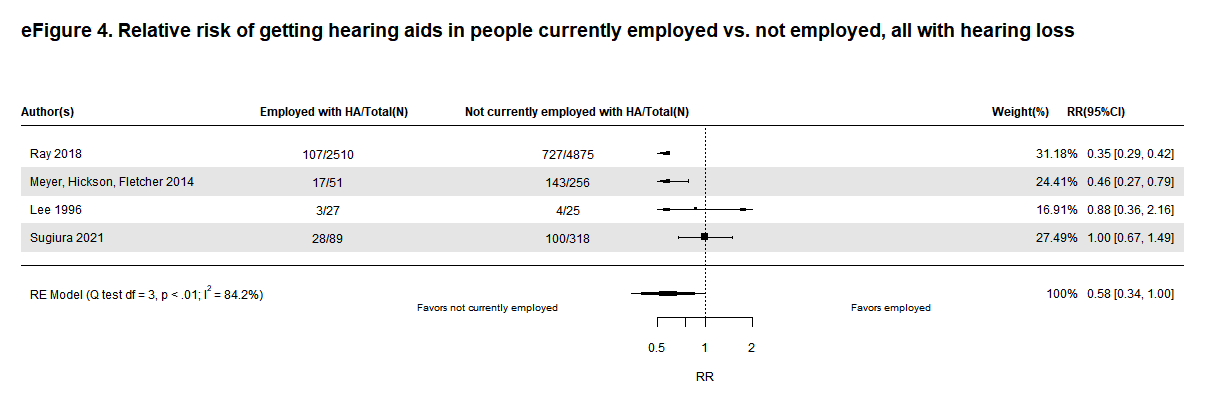

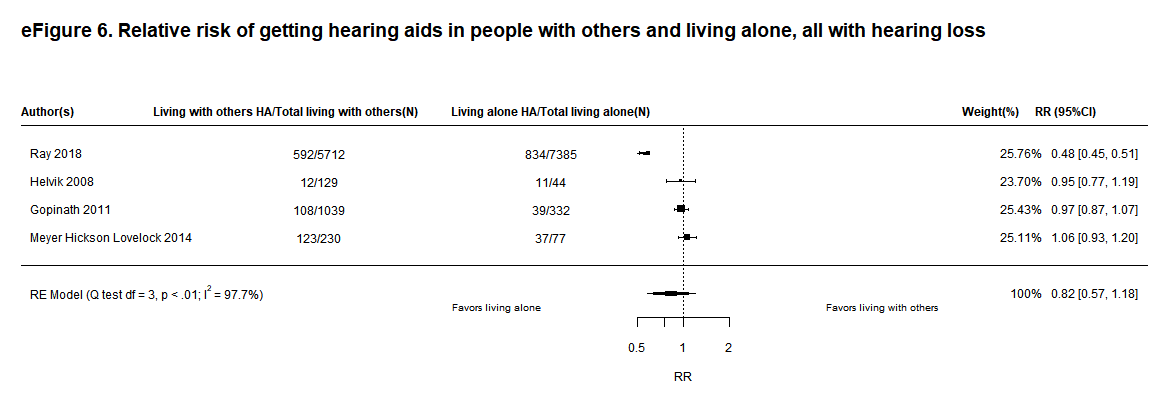

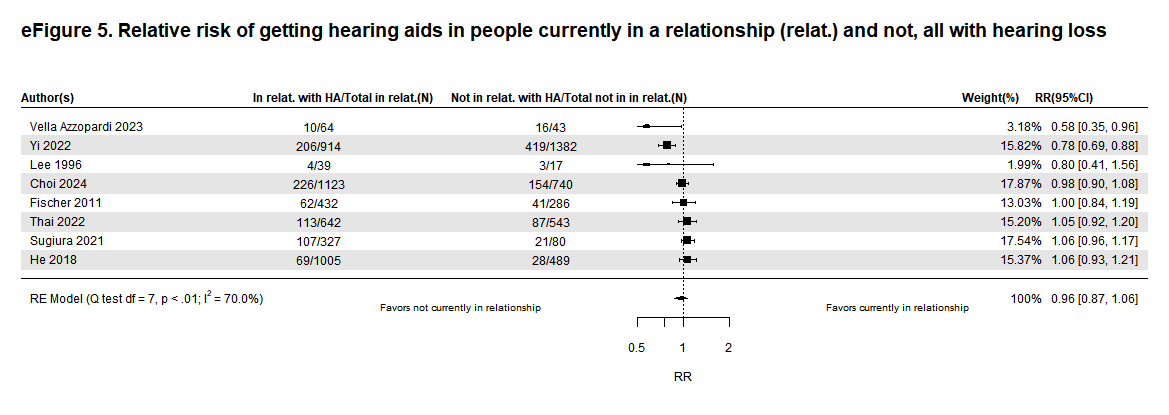

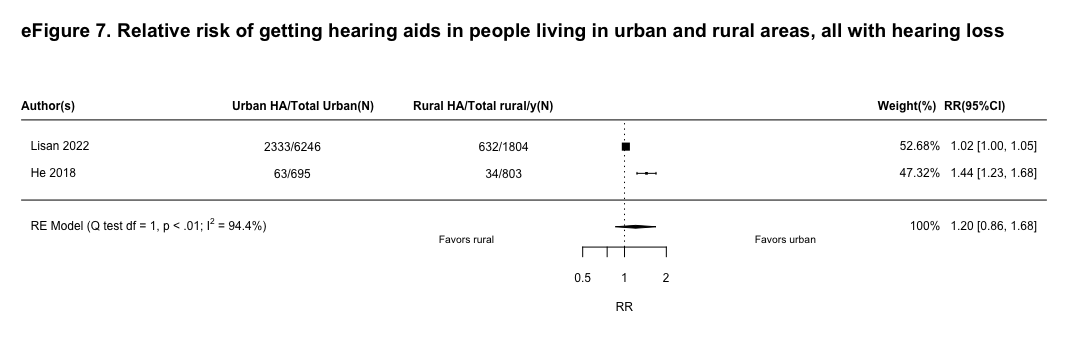

eTable 1: MOOSE Checklist for Meta-analyses of Observational Studies

| **Item No** | **Recommendation** | **Reported (Yes/No)** | **Reported on Page No** |
| --- | --- | --- | --- |
| Reporting of background should include | | | |
| 1 | Problem definition | Yes | 5 |
| 2 | Hypothesis statement | Yes | 6 |
| 3 | Description of study outcome(s) | Yes | 7 |
| 4 | Type of exposure or intervention used | Yes | 7 |
| 5 | Type of study designs used | Yes | 7 |
| 6 | Study population | Yes | 5-7 |
| Reporting of search strategy should include | | | |
| 7 | Qualifications of searchers (eg, librarians and investigators) | Yes | 6, 7 |
| 8 | Search strategy, including time period included in the synthesis and key words | Yes | 6, 7; supplementary |
| 9 | Effort to include all available studies, including contact with authors | Yes | 6, 7 |
| 10 | Databases and registries searched | Yes | 6, 7 |
| 11 | Search software used, name and version, including special features used (eg, explosion) | Yes | 6, 7 |
| 12 | Use of hand searching (eg, reference lists of obtained articles) | Yes | 7 |
| 13 | List of citations located and those excluded, including justification | Yes | eFigure 1 |
| 14 | Method of addressing articles published in languages other than English | Yes | 6 |
| 15 | Method of handling abstracts and unpublished studies | Yes | 6 |
| 16 | Description of any contact with authors | Yes | 7 |
| Reporting of methods | | | |
| 17 | Description of relevance or appropriateness of studies assembled for assessing the hypothesis to be tested | Yes | 6-7 |
| 18 | Rationale for the selection and coding of data (eg, sound clinical principles or convenience) | Yes | 6-7 |
| 19 | Documentation of how data were classified and coded (eg, multiple raters, blinding and interrater reliability) | Yes | 7 |
| 20 | Assessment of confounding (eg, comparability of cases and controls in studies where appropriate) | Yes | 7 |
| 21 | Assessment of study quality, including blinding of quality assessors, stratification or regression on possible predictors of study results | Yes | 8 |
| 22 | Assessment of heterogeneity | Yes | 9 |
| 23 | Description of statistical methods (eg, complete description of fixed or random effects models, justification of whether the chosen models account for predictors of study results, dose-response models, or cumulative meta-analysis) in sufficient detail to be replicated | Yes | 9 |
| 24 | Provision of appropriate tables and graphics | Yes | 25-46; supplementary |
| Reporting of results should include | | | |
| 25 | Graphic summarizing individual study estimates and overall estimates | Yes | Figures 1-5; supplementary |
| 26 | Table giving descriptive information for each study included | Yes | Table 1 |
| 27 | Results of sensitivity testing (eg, subgroup analysis) | Yes | 10-13 |
| 28 | Indication of statistical uncertainty of findings | Yes | 9-13 |
| Reporting of discussion should include | | | |
| 29 | Quantitative assessment of bias (eg, publication bias) | Yes | 14, 18 |
| 30 | Justification for exclusion (eg, exclusion of non-English language citations) | No | N/A |
| 31 | Assessment of quality of included studies | Yes | 18 |
| Reporting of conclusions should include | | | |
| 32 | Consideration of alternative explanations for observed results | Yes | 18 |
| 33 | Generalization of the conclusions (ie, appropriate for the data presented and within the domain of the literature review) | Yes | 17, 18 |
| 34 | Guidelines for future research | Yes | 18 |
| 35 | Disclosure of funding source | Yes | 19 |

*From*: Stroup DF, Berlin JA, Morton SC, et al, for the Meta-analysis Of Observational Studies in Epidemiology (MOOSE) Group. Meta-analysis of Observational Studies in Epidemiology. A Proposal for Reporting. *JAMA*. 2000;283(15):2008-2012. doi: 10.1001/jama.283.15.2008.

eTable 2: Search strategies

**Embase**

| Set | Search Statement |
| --- | --- |
| 1. | (hearing intervention or hearing interventions or hearing aid or hearing aids or hearing device or hearing devices or hearing treatment or listening device or listening devices or hearing unit or hearing units or hearing instrument or hearing system or hearing systems).mp. [mp=title, abstract, heading word, drug trade name, original title, device manufacturer, drug manufacturer, device trade name, keyword heading word, floating subheading word, candidate term word] |
| 2. | hearing aid/ or assistive listening device/ or digital hearing aid/ or master hearing aid/ |
| 3. | retrospective study/ |
| 4. | prospective study/ |
| 5. | longitudinal study/ |
| 6. | (retrospective adj (study or studies)).tw. |
| 7. | (Cohort adj (study or studies)).tw. |
| 8. | (prospective adj (study or studies)).tw. |
| 9. | (longitudinal adj (study or studies)).tw. |
| 10. | (epidemiologic$ adj (study or studies)).tw. |
| 11. | (observational adj (study or studies)).tw. |
| 12. | (follow up adj (study or studies)).tw. |
| 13. | 1 or 2 |
| 14. | 3 or 4 or 5 or 6 or 7 or 8 or 9 or 10 or 11 or 12 |
| 15. | 13 and 14 |
| 16. | (exp animal/ or animal.hw. or nonhuman/) not (exp human/ or human cell/ or (human or humans).ti.) |
| 17. | 15 not 16 |

**Medline**

| 1. | (hearing intervention or hearing interventions or hearing aid or hearing aids or hearing device or hearing devices or hearing treatment or listening device or listening devices or hearing unit or hearing units or hearing instrument or hearing system or hearing systems).mp. [mp=title, book title, abstract, original title, name of substance word, subject heading word, floating sub-heading word, keyword heading word, organism supplementary concept word, protocol supplementary concept word, rare disease supplementary concept word, unique identifier, synonyms, population supplementary concept word, anatomy supplementary concept word] |
| --- | --- |
| 2. | Hearing Aids/ |
| 3. | Retrospective Studies/ |
| 4. | Longitudinal Studies/ |
| 5. | Prospective Studies/ |
| 6. | retrospective.tw. |
| 7. | longitudinal.tw. |
| 8. | (Follow up adj (study or studies)).tw. |
| 9. | (observational adj (study or studies)).tw. |
| 10. | (cohort adj (study or studies)).tw. |
| 11. | epidemiologic studies/ |
| 12. | Cohort Studies/ |
| 13. | 1 or 2 |
| 14. | 3 or 4 or 5 or 6 or 7 or 8 or 9 or 10 or 11 or 12 |
| 15. | 13 and 14 |
| 16. | exp animals/ not humans.sh. |
| 17. | 15 not 16 |

**Web of Science**

\# Searches:

1: TS=(“hearing intervention” OR “hearing interventions” OR “hearing aid” OR “hearing aids” OR “hearing device” OR “hearing devices” OR “hearing treatment” OR “listening device” OR “listening devices” OR “hearing unit” OR “hearing units” OR “hearing instrument” OR “hearing system” OR “hearing systems”)

2: TS=("cohort stud*" OR "retrospective stud*" OR "prospective stud*" or "longitudinal stud*")

3: #2 AND #1
