## Supplemental 2 for "Equality in Hearing Aid Access: A Systematic Review and Meta-analysis"

**Supplement 2: Data Sharing Statement**

Data

Data available: No

Additional information:

Explanation of why data is not available: We have provided the search terms, and a list of included papers in the publication. If any additional information is needed, we will provide it upon request.
